# The genomic landscape of Acute Respiratory Distress Syndrome: a meta-analysis by information content of genome-wide studies of the host response

**DOI:** 10.1101/2024.02.13.24301089

**Authors:** Jonathan E Millar, Sara Clohisey-Hendry, Megan McMannus, Marie Zechner, Bo Wang, Nick Parkinson, Melissa Jungnickel, Nureen Mohamad Zaki, Erola Pairo-Castineira, Konrad Rawlik, Joshua Rogers, Clark D Russell, Lieuwe DJ Bos, Nuala J Meyer, Carolyn Calfee, Daniel F McAuley, Manu Shankar-Hari, J Kenneth Baillie

## Abstract

Acute respiratory distress syndrome (ARDS) is a clinically defined syndrome of acute hypoxaemic respiratory failure secondary to non-cardiogenic pulmonary oedema. It arises from a diverse set of triggers and encompasses marked biological heterogeneity, complicating efforts to develop effective therapies. An extensive body of recent work (including transcriptomics, proteomics, and genome-wide association studies) has sought to identify proteins/genes implicated in ARDS pathogenesis. These diverse studies have not been systematically collated and interpreted.

To solve this, we performed a systematic review and computational integration of existing omics data implicating host response pathways in ARDS pathogenesis. We identified 40 unbiased studies reporting associations, correlations, and other links with genes and single nucleotide polymorphisms (SNPs), from 6,856 ARDS patients.

We used meta-analysis by information content (MAIC) to integrate and evaluate these data, ranking over 7,000 genes and SNPs and weighting cumulative evidence for association. Functional enrichment of strongly-supported genes revealed cholesterol metabolism, endothelial dysfunction, innate immune activation and neutrophil degranulation as key processes. We identify 51 hub genes, most of which are potential therapeutic targets. To explore biological heterogeneity, we conducted a separate analysis of ARDS severity/outcomes, revealing distinct gene associations and tissue specificity. Our large-scale integration of existing omics data in ARDS enhances understanding of the genomic landscape by synthesising decades of data from diverse sources. The findings will help researchers refine hypotheses, select candidate genes for functional validation, and identify potential therapeutic targets and repurposing opportunities. Our study and the publicly available computational framework represent an open, evolving platform for interpretation of ARDS genomic data.

## Introduction

The acute respiratory distress syndrome (ARDS) is clinically defined as acute hypoxaemic respiratory failure due to non-cardiogenic pulmonary oedema^1^. It occurs following a variety of insults; pulmonary and extra-pulmonary. While this definition has been useful in identifying patients at risk of serious morbidity and death^2^, it overlooks the underlying biology and masks heterogeneity^3^. Arguably, this has contributed to limited success in developing therapeutics^4^. In contrast, a biological definition of ARDS may provide the lever necessary for future drug discovery^5^.

Functional genomics technologies enable hypothesis-free disease characterisation at unprecedented resolution. The emergence of coronavirus disease 2019 (COVID-19) has provided an opportunity to test genetic approaches to drug discovery in a homogeneous subset of ARDS patients. A notable success is the finding that baricitinib, a Janus kinase inhibitor, reduces mortality in patients hospitalised with COVID-19^6^. *A priori* support for baricitinib^7^ was greatly enhanced following the discovery of a causal link between elevated tyrosine kinase 2 (TYK2) expression and severe COVID-19 in genome-wide association studies (GWAS)^8^. The availability of comparable omics data for non-COVID ARDS is limited.

An unresolved challenge is how large omics data can be effectively exploited^9^. Specifically, how can we combine data from heterogeneous sources to derive new insights or recalibrate our understanding in the light of new data? We have proposed meta-analysis by information content (MAIC) as a data-driven, algorithmic, method for combining gene lists from diverse sources^10^. MAIC is agnostic to the quality or methodology of the sources and combines ranked or un-ranked gene sets by calculating weights for each list and gene, and iteratively updating them to converge on a ranked meta-list. We have successfully applied MAIC to host-genomics studies of influenza A^10^ and coronavirus infection^8,11^, and shown that it out-performs existing algorithms when combining ranked and un-ranked lists obtained from heterogeneous sources^12^.

In this work, we present a living meta-analysis by information content of ARDS host genomics studies. This serves as an open-source resource for gene prioritisation, functional genomics, and drug target discovery. An interactive interface can be accessed at https://baillielab.net/maic/ards, alongside a complementary R package.

## Results

### Systematic review

We first conducted a systematic review of existing genome-wide studies, which reported associations between genes, transcripts, or proteins and ARDS susceptibility, severity, survival, or phenotype. Our search yielded 8,937 unique citations (Fig. S1). We retrieved 74 articles for full-text evaluation and included 40 in our meta-analysis^13–52^. These 40 studies produced 44 unique gene lists (22 transcriptomic, 13 proteomic, and 9 based on genome-wide association studies (GWAS); see Table 1). Three studies reported results from multiple methodologies^33,38,44^, and several used more than one tissue type^18,21,32^. Excluding GWAS, 14 gene lists (40%) were derived from lung or airways samples, and 21 (60%) from blood. We could not retrieve one gene list^26^. No whole-genome sequencing GWAS were found, and only 36% (n=8) of transcriptomic lists used next-generation sequencing techniques. The earliest included study was published in 2004^18^, however, almost half (n=19, 47.5%) were published in the last 5 years.

**Table 1:**
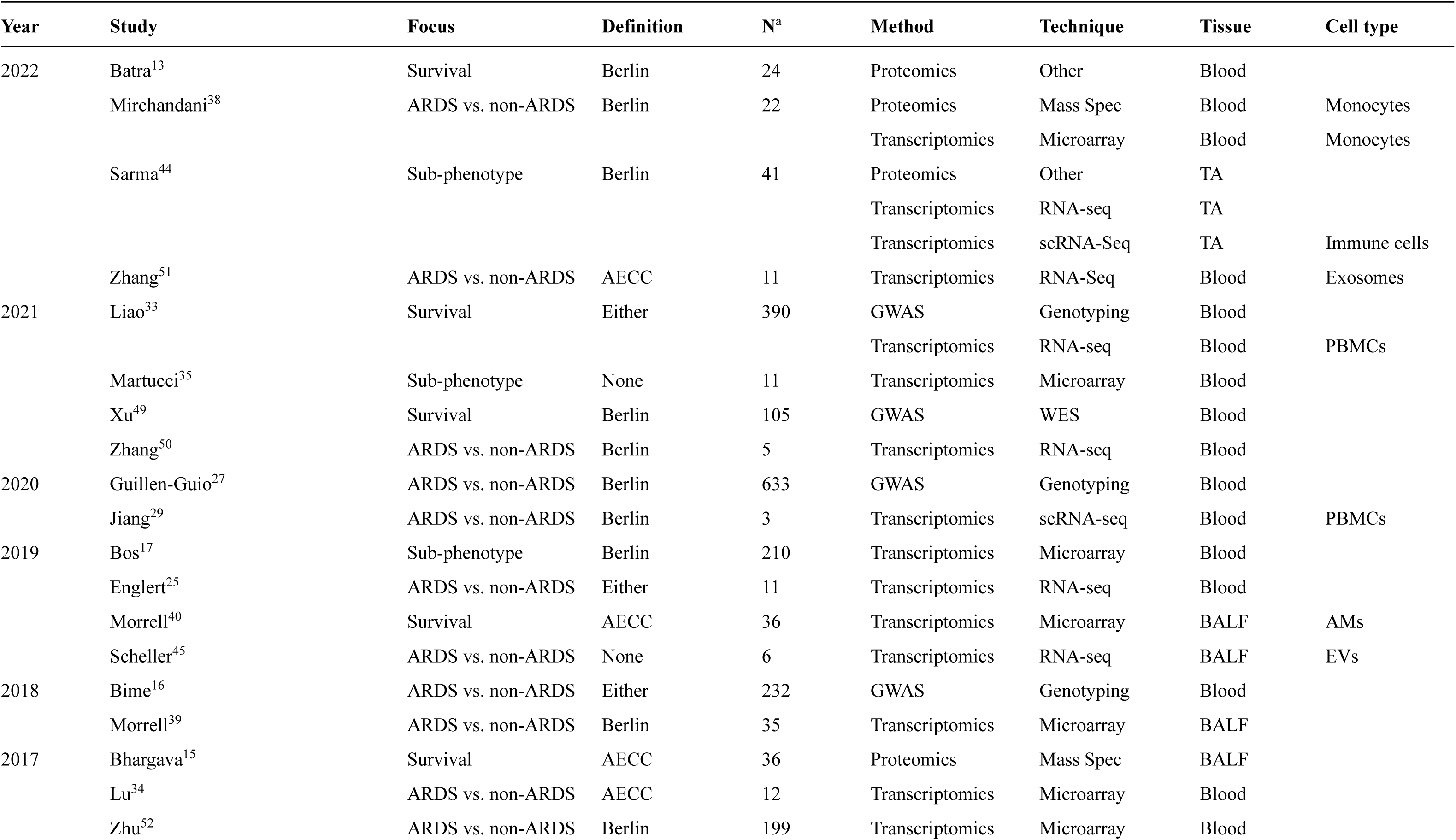
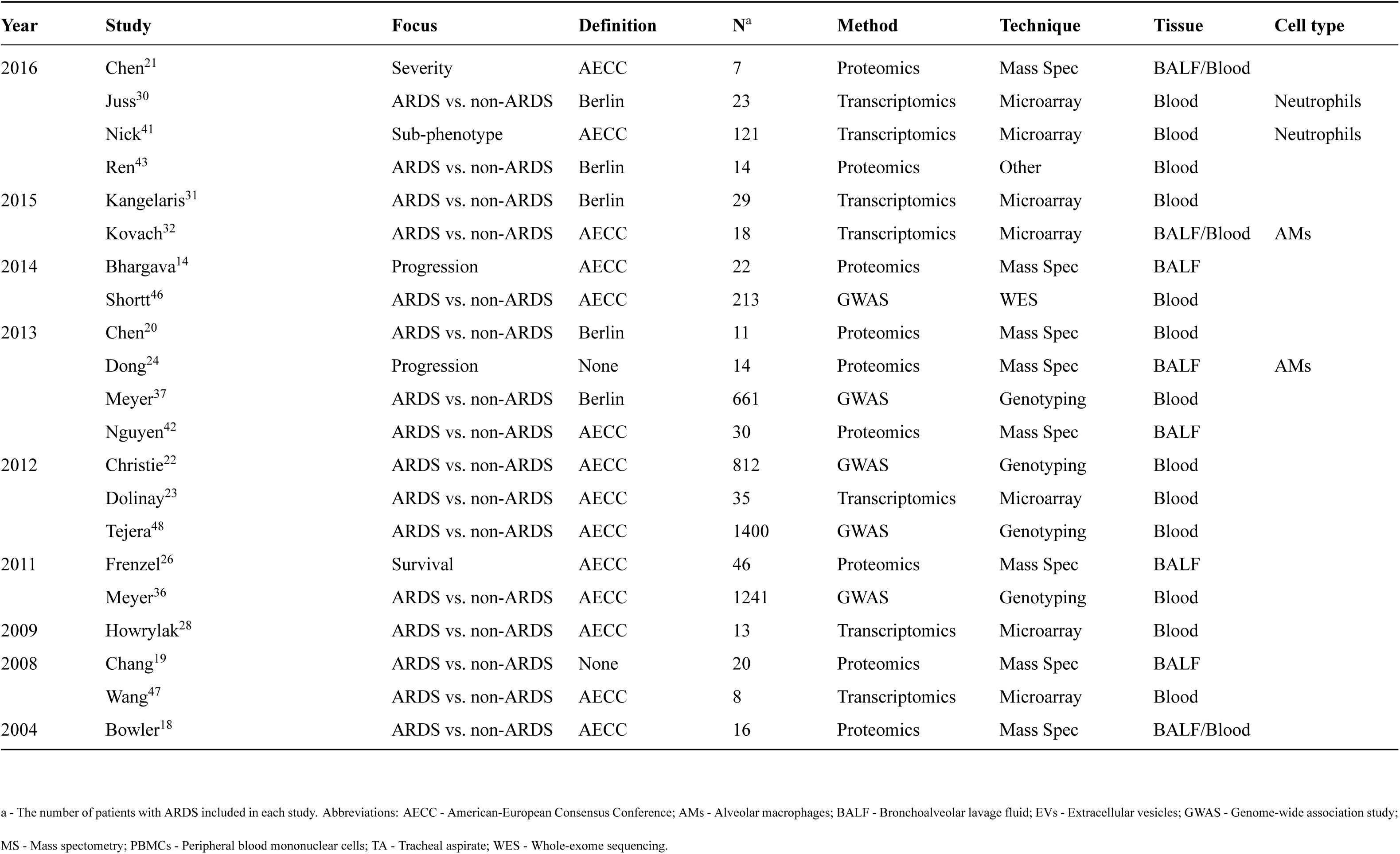
Summary of studies and gene lists included in the systematic review.

Most studies aimed to identify genes or proteins associated with ARDS susceptibility (n=27, 67.5%). The remainder examined associations with survival (n=6, 15%), sub-phenotype (n=4, 10%), disease progression (n=2, 5%), or severity (n=1, 2.5%). In total, studies included 6,856 patients with ARDS.

### Meta-analysis by information content (MAIC)

We analysed all 43 available gene lists using MAIC. Lists were categorised by method (i.e., GWAS, transcriptomics, and proteomics) and technique (e.g., RNA-seq, mass spectrometry; see Table 1). In total, we ranked 7,085 unique genes (or SNPs), with a median of 27 genes per gene list (range 1-4,954). The top 100 ranked genes are summarised in Figure 1. Most genes were found in a single category (n=5,866, 82.8%); only 157 (2.2%) were identified in ≥ 3 categories, with the maximum number of categories supporting a gene being 5 (Figure 1). Similarly, few genes (n=362, 5.1%) were identified by more than one method, with only *AKR1B10*, *HINT1*, *HSPG2*, *S100A11*, and *SLC18A1* present in transcriptomic, proteomic, and GWAS-based lists.

**Figure 1:**
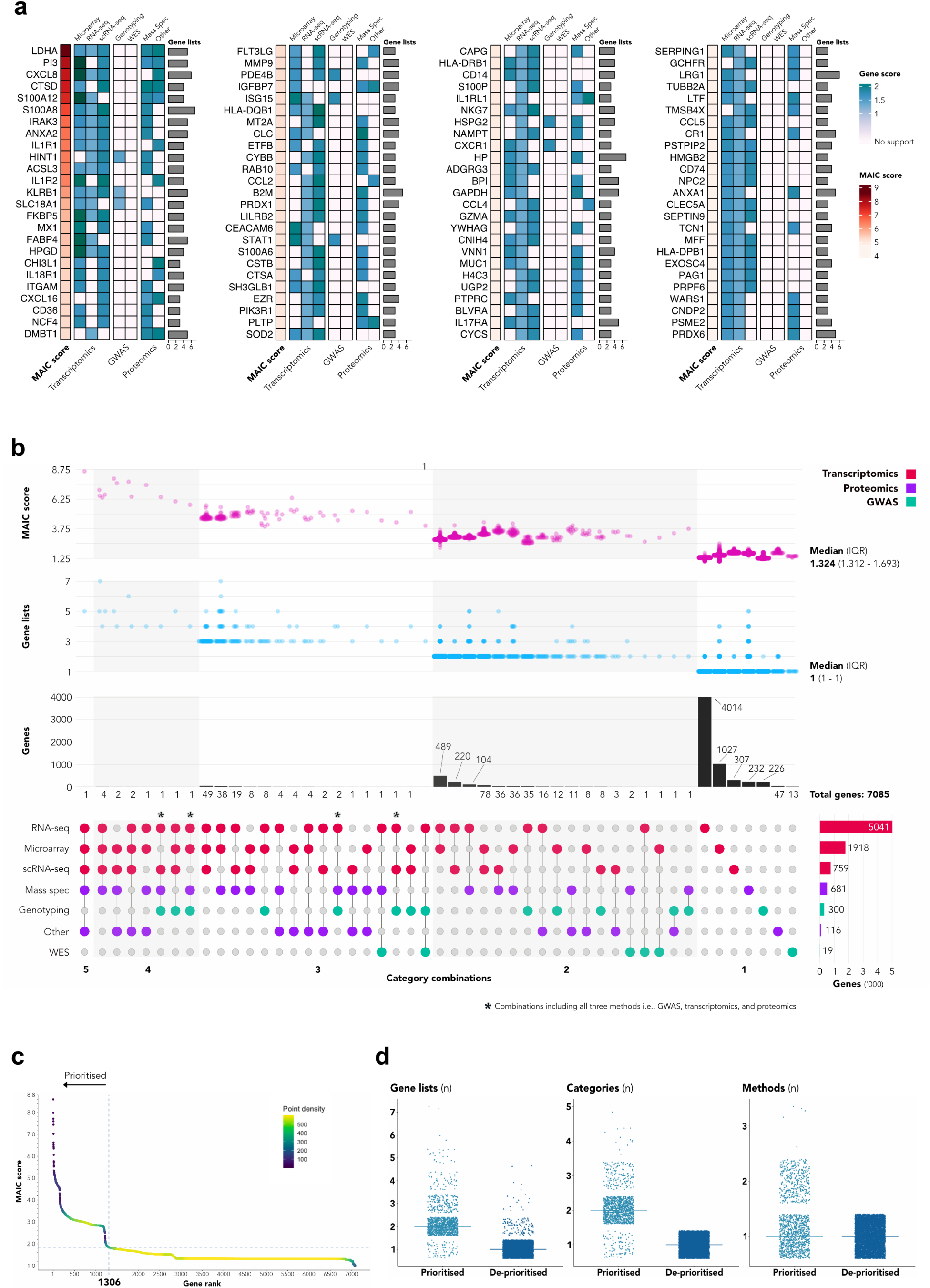
Meta-analysis by information content. (a) Heatmap of top 100 ranked genes showing MAIC score, highest score per category, and number of supporting lists. (b) UpSet plot of ranked genes showing total numbers for each category combination, MAIC score distribution, and supporting lists. (c) Gene prioritisation using the Unit Invariant Knee method. Intersection of lines identifies elbow point of best-fit curve. 1,306 genes in the upper left quadrant were prioritied. (d) Strip plots comparing number of lists, categories, and methods per gene between prioritised and deprioritised sets.

To prioritise genes for further investigation, we used the unit invariant knee method^53^ to identify the inflection point in the MAIC score curve. This prioritised 1,306 genes with scores above this point (Figure 1). These genes were more likely to be found in ≥ 2 lists or categories and by more than one method (Figure 1).

To assess the influence of individual lists, we calculated the total MAIC score (totMS), reflecting the sum of gene scores across each list (Fig. S2), and the contributing total MAIC score (ctotMS), measuring the sum of each lists gene scores which contribute to a gene’s overall MAIC score. To obtain relative values, we divided the totMS/ctotMS for each list by the total across all included lists. This demonstrated that only 10 lists (from 9 studies) contributed >1% by either metric (Tab. S1). Notably, the RNA-seq list from Sarma et al.^44^ accounts for >50%, a function of its length. To account for this, we normalised totMS/ctotMS by the number of genes per list; along with the proportion of replicated genes in each list, this provides an alternative perspective, with several proteomic studies ranking highly (Fig. S2).

### Comparison with existing ARDS sources and COVID-19

To place our meta-analysis results in context, we evaluated the overlap between the genes prioritised by MAIC and those from two established resources: BioLitMine^54^, using an ARDS MeSH search, and the ARDS Database of Genes^55^ (Fig. S3a and Fig. S3c). A search using BioLitMine, identified 271 ARDS-associated genes, of which 142 (52.4%) were present in our analysis. Almost half of the overlapping genes (n = 63, 44.4%) ranked within our prioritised set (Tab. S2).

After correcting for historical gene symbol aliases, we matched 4 additional genes from the BioLitMine search. A further 104 genes were supported by just a single publication (Fig. S3b). For each of the remaining 21 genes, we obtained the 100 most co-expressed genes using ARCHS4^56^ (returning data for 18) and assessed the overlap of these sets with the results of ARDS MAIC; two-thirds exhibited <50% overlap (Fig. S3b). Of the 239 genes catalogued in the ARDS Database of Genes, 177 (74.1%) were also found in our study. However, both sources contain gene associations which lack genome-wide support.

Finally, we compared the overlap between genes ranked by ARDS MAIC and those identified in a previous MAIC of the host response to coronaviruses^11^ (Fig. S3d). In total, 2,606 genes (36.8%) were shared, of which 143 were prioritised by both analyses (Fig. S3e).

### Tissue and cell-specific expression

While most gene lists were derived from blood sampling, most genes were identified in airways samples (n=5,847, 82.5%) (Fig. S4a). This was equally the case for the prioritised gene set, however the majority of these genes were also identified in blood samples (n=818, 62.6%) (Fig. S4b). Among genes uniquely identified in lists obtained from blood samples (n=1,238), almost three-quarters are known to be expressed in the lung (HPA scRNA-seq data, ≥ 5 normalised transcripts per million (nTPM)), with a quarter being highly-expressed (≥ 100 nTPM) (Fig. S4c).

For prioritised genes found in lists obtained from airways sampling, there was a wide variety of cell-specific expression (Fig. S4d). However, in the smaller set of prioritised genes identified solely in lists employing blood sampling, clusters of expression specific to neutrophils, T cells, and monocytes were evident (Fig. S4e). Cell-type specific gene enrichment analysis suggests innate immune as well as epithelial and endothelial cell types are enriched among genes identified in airways samples (Fig. S4f). However, enrichment of epithelial and endothelial cells was not evident for prioritised genes identified from blood sampling alone (Fig. S4g).

### Functional enrichment

Having identified a set of prioritised genes, we undertook several functional enrichment analyses. First, we performed over-representation analysis (ORA). In Reactome, 51 terms were significantly enriched (*P* < 0.001) (Figure 3). Not unexpectedly, neutrophil degranulation and several innate immune pathways (e.g., IL-10 signalling, interferon signalling, MHC II antigen presentation, TLR4 cascade) featured heavily. However, multiple pathways associated with cholesterol biology and metabolism (e.g., chylomicron assembly/remodelling, GLUT4 translocation, TP53 regulation of metabolic genes, insulin regulation) were also over-represented. Similarly, lipid and cholesterol metabolism, as well as hyperlipidaemia, were over-represented in KEGG and WikiPathways (Fig. S5a and Fig. S5b). In an enrichment analysis using the GWAS Catolog, the prioritised set of genes was associated with asthma (adult onset/time to onset), monocyte, lymphocyte, and eosinophil counts, aspartate aminotransferase levels, and levels of apolipoprotein A1 (Fig. S5d).

Next, we used the prioritised set of genes to create a protein-protein interaction (PPI) network. We graph-clustered this network, identifying 48 clusters with ≥ 5 members. Among the 10 largest clusters, we found programs associated with the proteaosome, cholesterol metabolism, interferon signalling, IL-6 signalling, and the complement cascade (Fig. S6). We then sought to use the PPI network to identify hub genes using an ensemble of topological methods. This analysis suggests 51 genes as being central to the wider network, which fall into clear clusters implicating plausible biological pathways, including innate immune cytokine signalling, and interferon response (Figure 2). The majority of hub genes (n=31, 61%) are currently druggable and include targets such as *IL-6*, *IL-17A*, *IL-18*, and *MAP3K14*.

**Figure 2:**
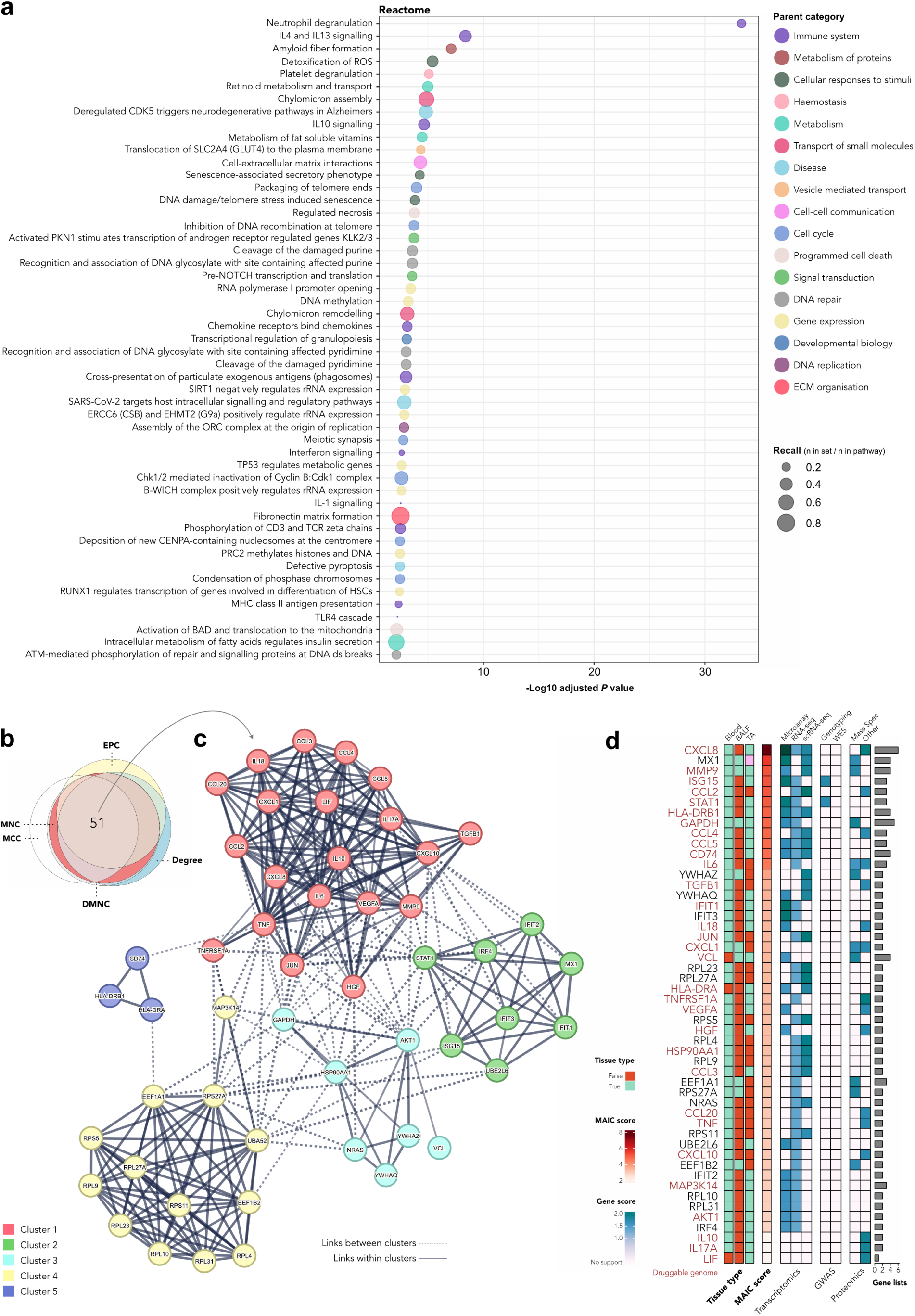
Functional enrichment of prioritised genes. (a) Significantly enriched Reactome terms (*P* < 0.01). Terms colored by parent class and size proportional to recall. (b) Euler diagram of the overlap of hub genes identified by five methods. MNC - Maximum Neighbourhood Component, MCC - Maximal Clique Centrality, DMNC - Density of MNC, EPC - Edge Percolated Component. (c) Protein-protein interaction (PPI) network of hub genes, clustered using MCL (Markov clustering). Clusters 1 and 2 contain canonical genes associated with innate immune cytokine signalling and interferon signalling respectively. Cluster 3 contains genes in the PI3K/AKT/mTOR pathway, which is an imporatant regulator of the cell cycle, and Cluster 4 contains ribosomal genes which are typically over-expressed during a stress response when protein synthesis increases. Cluster 5 contains genes required for antigen presentation through the MHC Class II pathway. (d). Heatmap of common hub genes displaying tissue type(s), MAIC score, highest category score, supporting lists, and presence in the DGIdb9 druggable genome (indicated in red).

### Sub-groups

To address the disparate range of study designs included in the overall analysis, we applied MAIC to key subsets of gene lists, with two different study desings: studies of ARDS versus non-ARDS controls (i.e. presence/absence of ARDS) (n=28) and studies of ARDS survival and severity (n=7) (Figure 3).

**Figure 3:**
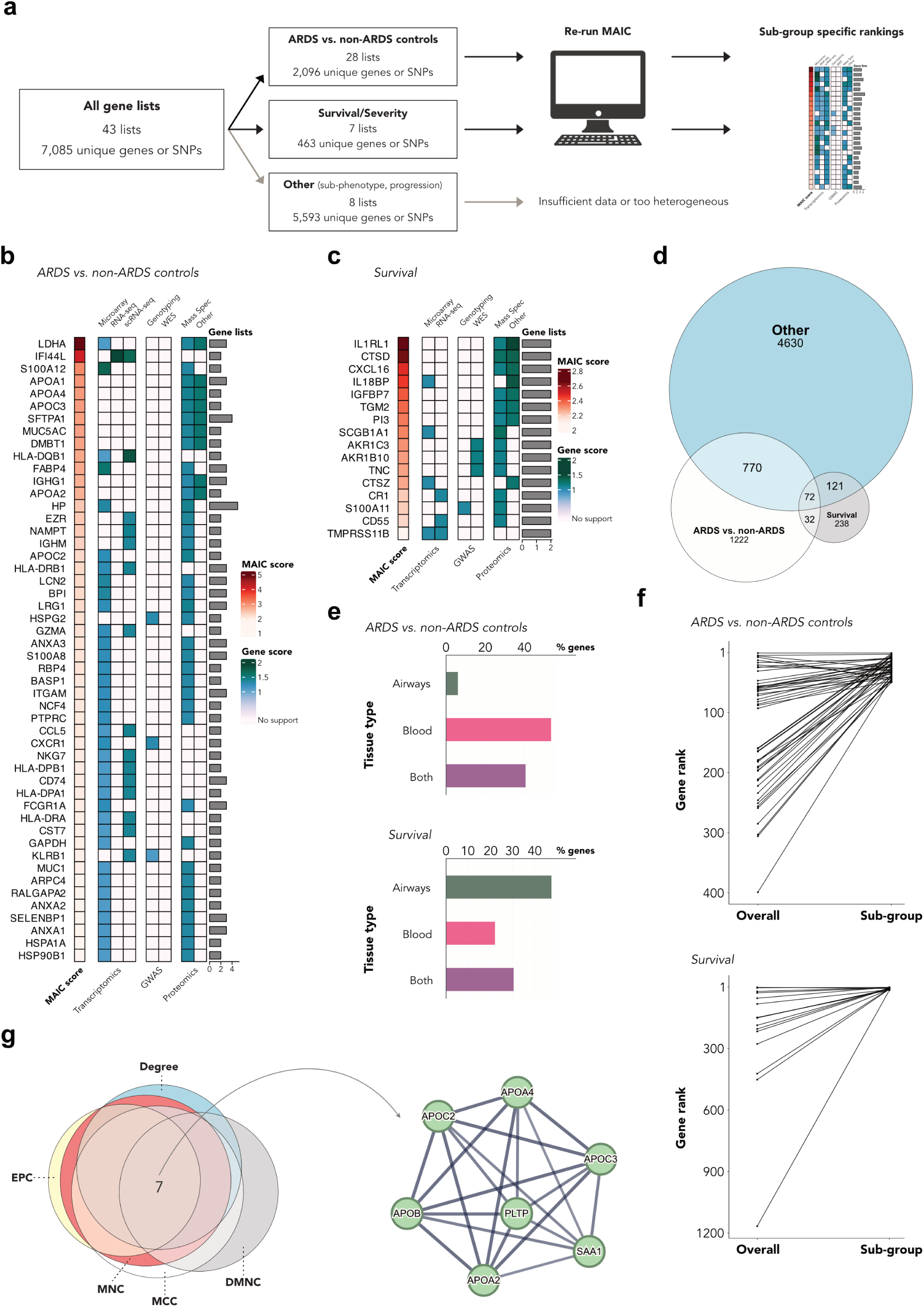
MAIC of sub-groups. (a) Schematic of ARDS MAIC sub-group analyses. (b) Heatmap of top 50 ranked genes in the ARDS vs. non-ARDS controls set showing MAIC score, highest score per category, and number of supporting lists. (c) Heatmap of 16 ranked genes in the survival set with multi-list support showing MAIC score, highest score per category, and number of supporting lists. (d) Euler diagram of gene overlap between the ARDS vs. non-ARDS controls and survival sets and the remainder of genes. (e) Bar plots of the tissue type in which genes are identified. (f) Slope plot comparing the ranks of ARDS vs. non-ARDS controls and survival prioritised genes with their ranks in the full iteration of ARDS MAIC. (g) Euler diagram of the overlap of hub genes identified by five methods. MNC - Maximum Neighbourhood Component, MCC - Maximal Clique Centrality, DMNC - Density of MNC, EPC - Edge Percolated Component and a protein-protein interaction (PPI) network of hub genes, clustered using the MCL - for ARDS vs. non-ARDS controls.

For ARDS vs. non-ARDS controls, there were 15 transcriptomic (54%), 7 GWAS (25%), and 6 proteomic studies (21%). Together, these studies included 5,713 patients with ARDS. MAIC ranked 2,096 genes (Figure 3). The majority of these (n=1,222; 58%) were unique to to this sub-group (Figure 3). Most were identified in blood, with a small fraction found solely in airways samples. The inflection point method prioritised the top-ranked 130 genes (Fig. S7a). In comparison to the BioLitMine search and the ARDS Database of Genes, 71/271 and 117/239 genes were found among this sub-group, respectively (Fig. S7b). A single study, a microarray-based transcriptomic list from Juss *et. al.*^30^, contributed the largest total MAIC score in this analysis (Tab. S3). ORA using Reactome, KEGG, and WikiPathways identified 25 significantly enriched pathways, including multiple terms related to cholesterol metabolism and glycolysis (Figure 4). A consensus of topological models identified 7 hub genes within a PPI network of prioritised genes. These genes cluster in a single grouping related to cholesterol metabolism (Figure 3).

**Figure 4:**
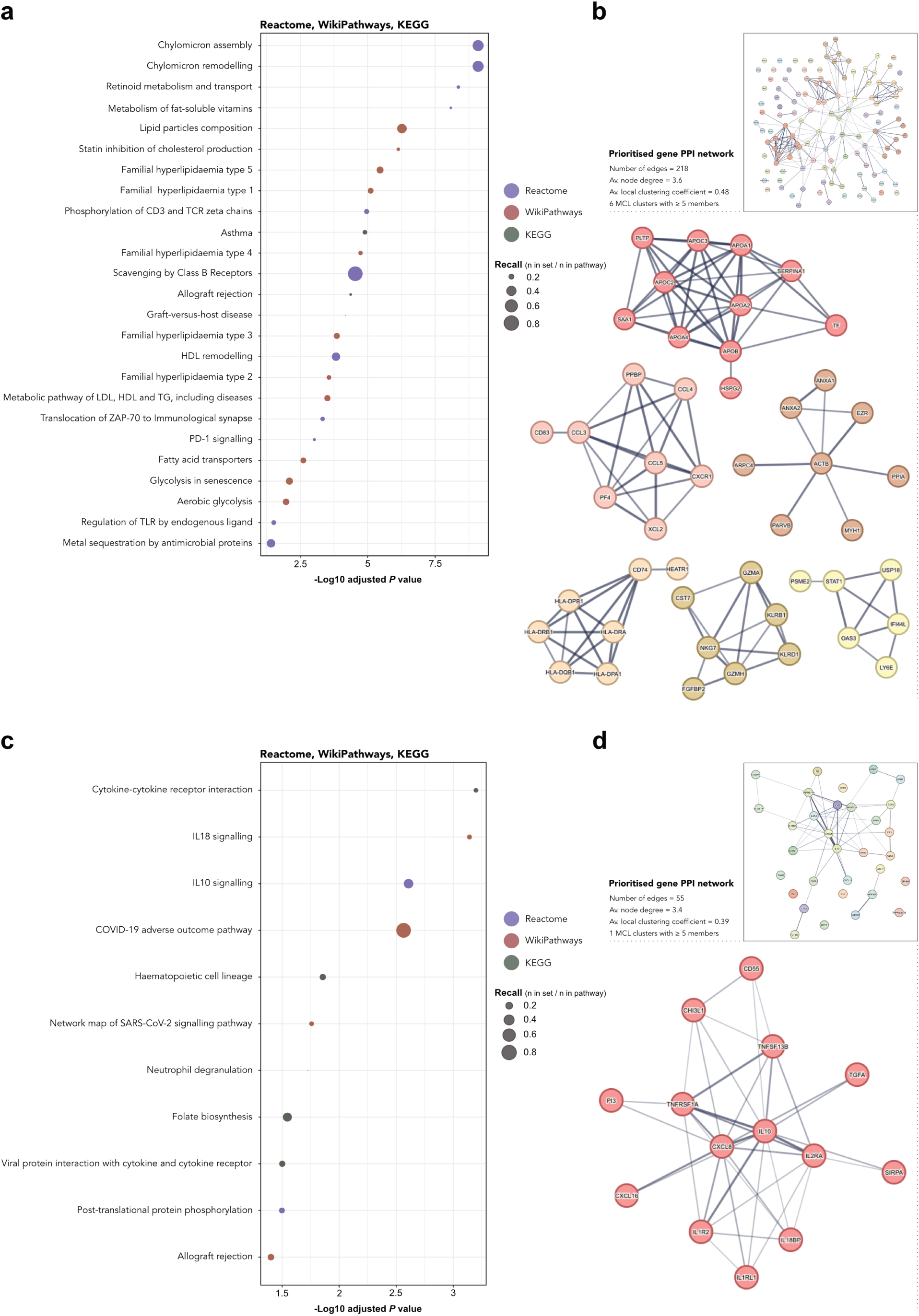
Sub-group functional enrichment. (a) Significantly enriched Reactome, WikiPathways, and KEGG terms (*P* < 0.01) for prioritised genes in the ARDS vs. non-ARDS controls sub-group. Terms are coloured by pathway and size is proportional to recall. (b) A protein-protein interaction network of prioritsed genes in the ARDS vs. non-ARDS controls cohort and graph-based clusters with ≥ 5 members. (c) Significantly enriched Reactome, WikiPathways, and KEGG terms (*P* < 0.01) for prioritised genes in the survival sub-group. Terms are coloured by pathway and size is proportional to recall. (d) A protein-protein interaction network of prioritsed genes in the survival cohort and graphbased clusters with ≥ 5 members.

In the survival/severity analysis, there were 8 gene lists, consisting of 3 transcriptomic lists (37.5%), 3 proteomic lists (37.5%), and 2 very small GWAS (25%). Together, these studies included 644 patients with ARDS. MAIC ranked 463 genes (Figure 3). Approximately half of these (n=238, 51%) were unique to survival-based lists. In contrast to the ARDS vs. non-ARDS analysis, most survival genes were found in airways samples. Thirty-three genes were prioritised (Fig. S7d). In total, 32/271 of the BioLitMine ARDS-associated genes and 23/239 of the ARDS Database of Genes genes were found among the ARDS MAIC survival set (Fig. S7e). The proteomic and transcriptomic lists from Bhargava *et.al*^15^ and Morrell *et. al*^40^ each contributed approximately 30% of the summed ctotMS of all included gene lists (Tab. S4). IL-10 and IL-18 signalling pathways were both significantly enriched in ORA (Figure 4). Graphbased (MCL) clustering of the prioritised set of survival genes identified a single large cluster of immune-related genes including, *IL-10*, *CXCL8*, *TNFRSF1A*, and *IL2RA* (Figure 4).

## Discussion

Our large-scale meta-analysis of the genomic landscape of ARDS prioritises 1,306 genes. Using wide inclusion criteria, we capture a diverse range of study designs and methods; the subsequent application of MAIC downgrades noisy, irrelevant, or low-quality information. These results have three main applications. First, they can be used to better understand the pathobiology of ARDS, providing a resource to prioritise future *in-vitro* and *in-vivo* studies and permitting comparisons between important sub-groups. Second, they prioritise therapeutic targets, serving as a resource against which novel and repurposed treatments can be screened. Third, they serve as a base for quantifying the novelty or additive nature of future ARDS studies using high-throughput technologies.

Our review included 40 studies taking a genome/proteome-wide approach with a variety of aims and methods. The rate at which this form of study is being published is increasing; half of all studies in the last 5 years and a quarter since 2020. Similarly, there were few studies which employed next-generation sequencing (NGS) techniques or equivalent, and only two single-cell RNA-seq studies. A partial explanation may be the emergence of COVID-19, which is likely to have consumed the attention of many research teams active in this field. We anticipate that an increasing number of non-COVID ARDS single-cell and NGS studies will emerge in the coming years. This reinforces the requirement for methods capable of meta-analysing multi-omic data^57^. Less obviously. A minority of studies have sampled the lung in ARDS, with only four examining the bulk transcriptome in the distal airspace. Reliance on information derived from blood samples may present a skewed picture of the pathobiology of ARDS and may be a missed opportunity to identify novel targets in the lung^58^.

A key advantage of the MAIC approach is its ability to integrate diverse data sources and deprioritise irrelevant information or noise. Traditional methods of gene list meta-analysis rely on simple vote counting or robust rank aggregation^59^. Instead, MAIC applies a data-derived weighting to each gene list, allows the investigator to define granular categorisation (preventing any one particular method from overwhelming the analysis), and permits the inclusion of both ranked and unranked lists. For data structures common in biological research (high noise, heterogeneity between studies, large input lists), MAIC outperformed other methods in a comprehensive simulation^12^. We have previously used MAIC to identify anti-viral genes in response to influenza A infection^10^ and Covid-19^11^.

Our results reinforce existing associations and reveal some new insights. The functional prominence of innate immunity and cytokine signalling - in particular neutrophil-related activity - is well described in the ARDS literature^60^, as is the high ranking of genes such as *CXCL8*^61^, *IL-18*^62^, *MMP9*^63^, and *MUC1*^64^. However, we also identify several genes that are consistently highly ranked in multiple studies, but have not been extensively discussed in the literature. Histidine triad nucleotide binding protein 1 (*HINT1*), ranked 10^th^ in our MAIC analysis, is one of only 5 genes to have support from GWAS, transcriptomics, and proteomics methods. To our knowledge, no role for *HINT1* has previously been suggested in ARDS^65^. However, *HINT1* has been implicated in T-cell response^66^, immunoregulation^67^, and apoptosis^65^. There is significant enrichment of cholesterol uptake, efflux, and esterification pathways among prioritised genes^68,69^. Stratification by sub-group revealed a tight cluster of genes important in cholesterol metabolism at the hub of those prioritised in ARDS vs. non-ARDS controls. This is of considerable therapeutic relevance given the potential role of drugs targeting this pathway in ARDS therapy^70,71^.

Multiple distinct pathways were identified in the setting of ARDS vs. non-ARDS controls, including type I interferon signalling^72^, MHC class II antigen presentation^73^, cell-cell adhesion^74^, and natural killer cell cytotoxicity^75^. In contrast, genes prioritised in our severity/outcome analysis are functionally more homogeneous and related to cytokine signalling, in particular IL-10 and IL-18 signalling. Our approach cannot determine whether this indicates a real difference between pathways active in ARDS (from the ARDS vs. non-ARDS analysis) and pathways associated with severity/survival, or an imbalance of study design or a of lack statistical power to detect some pathways in the severity/survival studies that have been conducted. We provide an open platform and associated tools to enable deeper mining of the output, allowing others to re-analyse the data based on alternative sub-group divisions or to integrate unseen information (https://github.com/baillielab/ARDSMAICr).In future, the addition of data from new technologies, and in greater scale and precision from existing technologies, is expected to substantially improve this analysis. For this reason, we consider this report to be the beginning of an ongoing, community-led multi-omic data integration.

Our approach has limitations. The majority of the original studies do not have designs that support causal inference, so we make no attempt to determine causality. Different methodologies, such as large-scale GWAS/meta-analysis and genome-wide summary Mendelian randomisation, may support causal inference in future. At present the available GWAS data is underpowered for this purpose. We purposefully excluded single-gene or candidate genetics studies. In the case of a gene with extensive evidence from the latter, our methodology may underestimate its association with ARDS. However, these study designs are subject to other limitations, such as publication and investigator biases and spurious associations arising from underpowered studies^76^. The limitations of the available data prevented us from accounting for direction of expression or effect. For a given gene, if the direction of expression differs between studies, we may therefore overestimate the strength of evidence associated with that gene. This also limits the scope of functional enrichment analyses which can be performed. Finally, the paucity of available data, and in particular the limited number of studies reporting data from ARDS subtypes or single-cell transcriptomics (or proteomics) studies, is an unavoidable limitation. It is likely that many pathological perturbations are highly cell-type and -state specific, and specific to distinct underlying disease process, which may not be apparent in bulk analyses of heterogeneous tissues identified using syndromic definitions^77^.

Our study provides a first step in systematically integrating decades of work in ARDS. Our results implicate potential therapeutic targets including interferon signalling and cholesterol metabolism dysregulation. Enrichment patterns and sub-group differences also give clues to genomic drivers of susceptibility, outcomes, and mortality. We show that combining existing data reveals new insights that were not observed in the original studies, and provide a framework for a living summary of the genomic landscape of ARDS.

## Methods

The systematic review and meta-analysis protocol was registered with the International Prospective Register of Systematic Reviews (PROSPERO; CRD42022306270). The review is reported in compliance with the Preferred Reporting Items for Systematic Reviews and Meta-Analyses (PRISMA) guidelines^78^.

### Search strategy and selection criteria

A detailed description of our search strategy and eligibility criteria is provided in the Supplementary Methods. Briefly, we searched MEDLINE, Embase, bioRxiv, medRxiv, the ARDS Database of Genes^55^, and the NCBI Gene Expression Omnibus from inception to April 1^st^, 2023 without language restrictions. We also performed single-level backwards and forwards citation searches using SpiderCite^79^ and hand-searched recent review articles^80–83^.

We included human genome-wide studies reporting associations between genes, transcripts, or proteins and ARDS susceptibility, severity, survival, or phenotype, accepting any contemporaneous ARDS definition. We excluded paediatric studies (age < 18 years), animal studies, *in-vitro* human ARDS models, candidate *in-vivo* or *in-vitro studies* (< 50 genes/proteins), candidate gene associations, and studies with < 5 patients per arm (except scRNA-seq).

### Outcomes

We retrieved ranked lists of genes associated with the ARDS host response, preferring measures of significance and adjusted *P* values over raw *P* values when multiple ranking measures were used. We obtained both summary lists (all implicated genes) and author-defined subgroup lists. To combine subgroup lists into summary lists, we took the minimum *P* value or maximum effect size. We excluded genes below the author-defined threshold for significance/effect magnitude. If unavailable, we excluded genes with *P* > 0.05, z-score < 1.96, or log fold change < 1.5.

### Study selection and data extraction

Article titles and abstracts from our search were stored in Zotero v6.0-beta (Corporation for Digital Scholarship, United States). Titles were initially screened by one author using Screenatron^79^. Two authors then independently screened abstracts against eligibility criteria, with a third resolving inconsistencies. Full texts and supplements of eligible studies were retrieved and inclusion adjudicated by consensus.

Data were extracted by one author and cross-checked by a second. Gene, transcript, or protein identifiers were mapped to HGNC symbols or Ensembl/RefSeq equivalents if no HGNC symbol was available. Unannotated SNPs were searched in NCBI dbSNP. miRBase (University of Manchester, United Kingdom) provided miRNA symbols. For microarray probes without symbols, we used the DAVID Gene Accession Conversion tool (Laboratory of Human Retrovirology and Immunoinformatics, Frederick National Laboratory for Cancer Research, United States) to map them to HGNC symbols. We extracted information relating to study design, methodology, tissue/cell type, demographics, ARDS aetiology, risk factors, severity, and outcomes.

### Meta-analysis by information content (MAIC)

The MAIC algorithm has been described in detail^8,10,11,84^. Full documentation and the source code are available at https://github.com/baillielab/maic. Briefly, MAIC combines ranked and unranked lists of related named entities, such as genes, from heterogeneous experimental categories, without prior regard to the quality of each source. The algorithm makes four key assumptions; (1) genes associated with ARDS exist as true positives, (2) a gene is more likely to be a true positive if it is found in more than one source, (3) the probability of being a true positive is enhanced if the gene appears in a list that contains a higher proportion of replicated genes, and (4) the probability is further enhanced if it is found in more than one category of experiment. Based on these assumptions, MAIC compares lists with each other, forming a weighting for each source based on its information content, which is then used to calculate a score for each gene. The output is a ranked list summarizing the total information supporting each gene’s association with ARDS. We have shown MAIC outperforms available algorithms, especially with ranked and unranked heterogeneous data^84^.

As our primary analysis, we performed MAIC on all summary gene lists, regardless of study focus. Lists were assigned categories based on their methodology and experimental technique: genome-wide association study (GWAS) - genotyping, GWAS - whole exome sequencing, transcriptomics - microarray, transcriptomics - RNA-sequencing (RNA-seq), transcriptomics - single cell RNA-seq (scRNA-seq), proteomics - mass spectometry, and proteomics - other. For secondary analyses, we performed MAIC on subsets of lists based on study focus (i.e., susceptibility to ARDS or survival/severity).

In secondary analyses, we repeated this pipeline for gene lists arising from studies in which the focus was ARDS vs. non-ARDS controls or ARDS survival/severity.

For each MAIC iteration, we prioritised genes with sufficient evidential support for further study (i.e., the gene set before which information content diminished such that there was little/no corroboration for the remainder’s ARDS association). We used the unit invariant knee method^53,85^ to identify the elbow point in the best-fit curve of MAIC scores. Genes with values above this point were prioritized for downstream analyses.

### ARDS literature and SARS-CoV-2 associations

We used BioLitMine^54^ to query the NCBI Gene database for genes associated with the Medical Subject Heading (MeSH) term “Respiratory Distress Syndrome, Acute”, generating a list of genes and publications. We descriptively compared the overlap between this list and the MAIC-ranked gene list. Similar comparisons were made between the ARDS MAIC results and the gene set in the ARDS Database of Genes^55^ and a prior MAIC of SARS-CoV-2 host genomics^11^.

### Tissue expression and enrichment

Transcript and protein expression data for genes included in ARDS MAIC were retrieved from the Human Protein Atlas (HPA, version 21.0)^86^. We investigated mRNA expression in a consensus scRNA-seq dataset of 81 cells from 31 sources (https://www.proteinatlas.org/about/assays+annotation#singlecell_rna) and in the HPA RNA-seq blood dataset^87^, containing expression levels in 18 immune cell types and total peripheral blood mononuclear cells. To investigate protein expression, we retrieved tissue-specific expression scores from the HPA^88^. We conducted cell-type specific enrichment analysis using WebCSEA^89^ and extracted the top 20 general cell types for each query.

### Functional enrichment

We performed functional enrichment of genes against the universe of all annotated genes using g:Profiler^90^. The following data sources were used; Kyoto Encyclopaedia of Genes and Genomes (KEGG)^91^, Reactome^92^, WikiPathways^93^, and Gene Ontology^94^. Multiple testing was corrected for using the g:SCS algorithm^90^, with a threshold of *P* < 0.01. Input lists were ordered by MAIC score were appropriate. In the case of GO cellular component terms, we used the REVIGO tool to perform multi-dimensional scaling of the matrix of all pairwise semantic similarities^95^. Enrichment was also performed against the National Human Genome Research Institute GWAS Catalog^96^ using the Enrichr web-interface^97^. Protein-protein interaction enrichment was performed using STRING v11^98^. We included all possible interaction sources but specified a minimum interaction score of 0.7. We used the the whole annotated genome as the statistical background. The MCL (Markov Clustering) algorithm[PMID: 22144159] was applied to the resulting network with an inflation parameter of 3. Clusters were annotated by hand having considered enrichment against KEGG, Reactome, and WikiPathways. To identify hub genes within the PPI network, we used cytoHubba^99^ and Cytoscape^100^. The highest ranked genes by Maximum Neighbourhood Component (MNC), Maximal Clique Centrality (MCC), Density of MNC (DMNC), Edge Percolated Component (EPC), and node degree were retrieved. The intersecting genes of these methods were deemed hub genes. Hub genes were searched for in the Drug Gene Interaction Database^101^ to identify if they were present in the druggable genome. The Drug Gene Interaction Database (DGIdb) was queried for each ranked gene^102^.

### Software and code availability

MAIC is implemented in Python v3.9.7 (Python Software Foundation, Wilmington, United States). All other analyses were performed with R v4.2.2 (R Core Team, R Foundation for Statistical Computing, Vienna, Austria). Code required to reproduce the analyses is available at https://github.com/JonathanEMillar/ards_maic_analysis. An R package (ARDSMAICR) containing the data used in this manuscript and several functions helpful in analyses is available at https://github.com/baillielab/ARDSMAICr.

## Data Availability

All data produced are available online at https://github.com/JonathanEMillar/ards_maic_manuscript/tree/main/data

## Supplementary Material

### Supplementary Methods

#### Search Strategy

We used the following strategy to search MEDLINE and a direct translation to search Embase.

**1** exp Respiratory Distress Syndrome, Adult/

**2** “acute lung injury*“.ti,ab,kf,kw

**3** 1 OR 2

**4** “gene*“.mp

**5** “genome*“.mp

**6** “transcript*“.mp

**7** “protein*“.mp

**8** 4 OR 5 OR 6 OR 7

**9** 3 AND 8

**10** (“COVID-19*” OR “COVID19*” OR “COVID-2019*” OR “covid”).ti,ab,kf,kw

**11** (“SARS-CoV-2*” OR “SARSCov-2*” OR “SARSCoV2*” OR “SARS-CoV2”).ti,sh,kf,kw

**12** (“2019-nCoV*” OR “2019nCoV*” OR “19-nCoV*” OR “19nCoV*” OR “nCoV2019*” OR “nCoV-2019*” OR “nCoV19*” OR “nCoV-19*“).ti,ab,kf,kw

**13** 10 OR 11 OR 12

**14** 9 NOT 13

**15** Letter.pt OR Conference Abstract.pt OR Conference Paper.pt OR Conference Review.pt OR Editorial.pt OR Erratum.pt OR Review.pt OR Note.pt OR Tombstone.pt

**16** 14 NOT 15

**17** exp *adolescence/ or exp *adolescent/ or exp *child/ or exp *childhood disease/ or exp *infant disease/ or (adolescen* or babies or baby or boy? or boyfriend or boyhood or girlfriend or girlhood or child or child* or child*3 or children* or girl? or infan* or juvenil* or juvenile* or kid? or minors or minors* or neonat* or neo-nat* or newborn* or new-born* or paediatric* or peadiatric* or pediatric* or perinat* or preschool* or puber* or pubescen* or school* or teen* or toddler? or underage? or under-age? or youth*).ti,kw

**18** 16 NOT 17

**19** ((exp animal/ or nonhuman/) NOT exp human/)

**20** 18 NOT 19

**21** limit 20 to yr=“1967-Current”

#### Inclusion criteria

Inclusion:

- Human studies: *in-vivo* or *in-vitro*
- Adults (age ≥ 18 years)
- Acute Respiratory Distress Syndrome (ARDS)

– by any contemporaneous definition
- Accepted methodologies:

– CRISPR screen
– RNAi screen
– Protein-protein interaction study
– Host proteins incorporated into virion or virus-like particle
– Genome wide association study
– Transcriptomic study
– Proteomic study

Exclusion:

- Children (age < 18 years)
- Animal studies
- Meta-analyses, *in-silico* analyses, or re-analysis of previously published data
- Excluded methodologies:

– *In-vitro* human studies simulating ARDS
– Candidate *in-vivo* or *in-vitro* transcriptomic or proteomic studies (defined as those investigating < 50 genes)
– Candidate gene association studies
– Studies including fewer than 5 individuals in either the control or ARDS arm

### Supplementary Results

**Supplementary-Figure 1:**
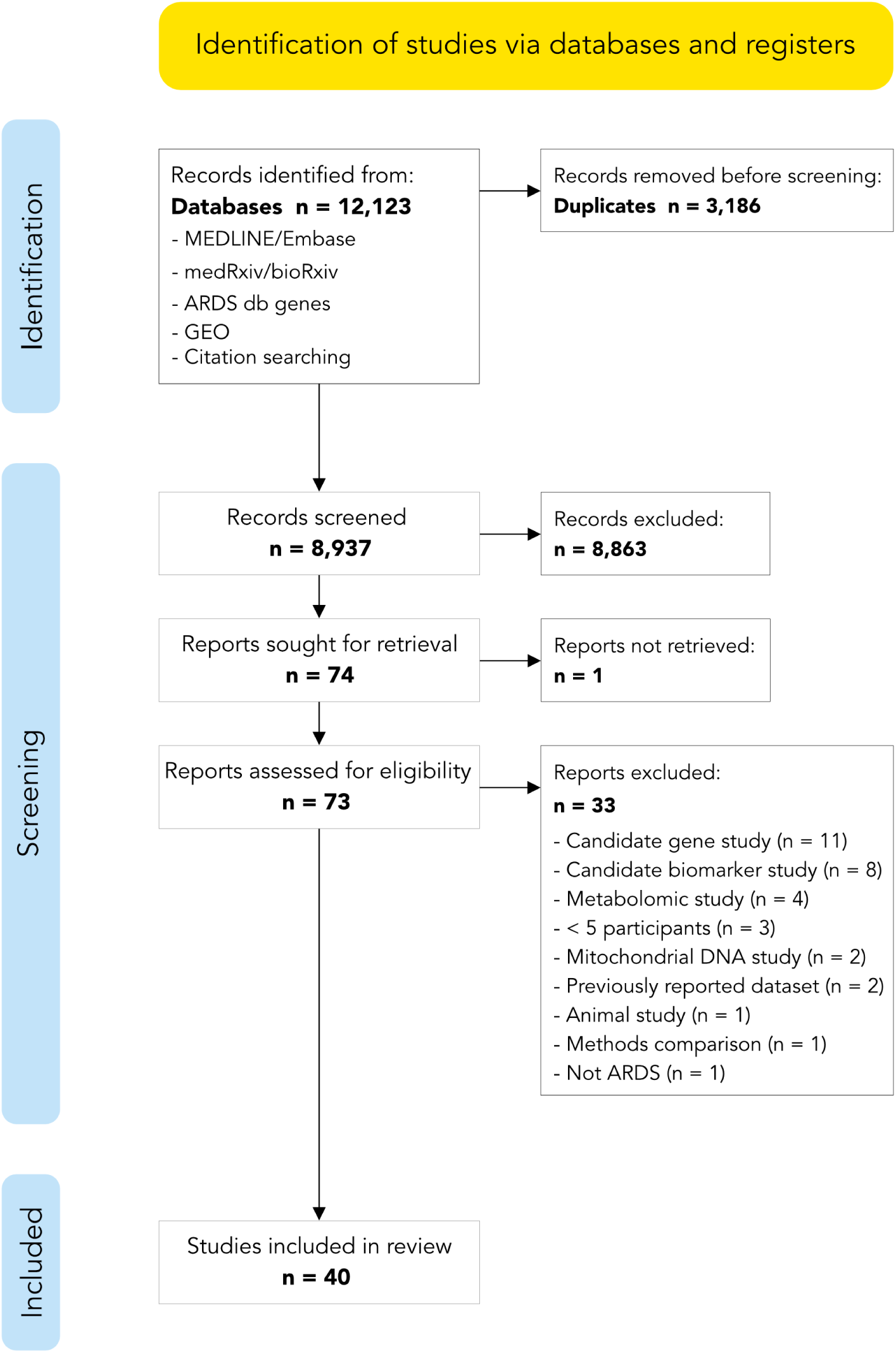
Systematic review inclusion diagram. Abbreviations: db - data base; GEO - NCBI Gene Expression Omnibus.

**Supplementary-Figure 2:**
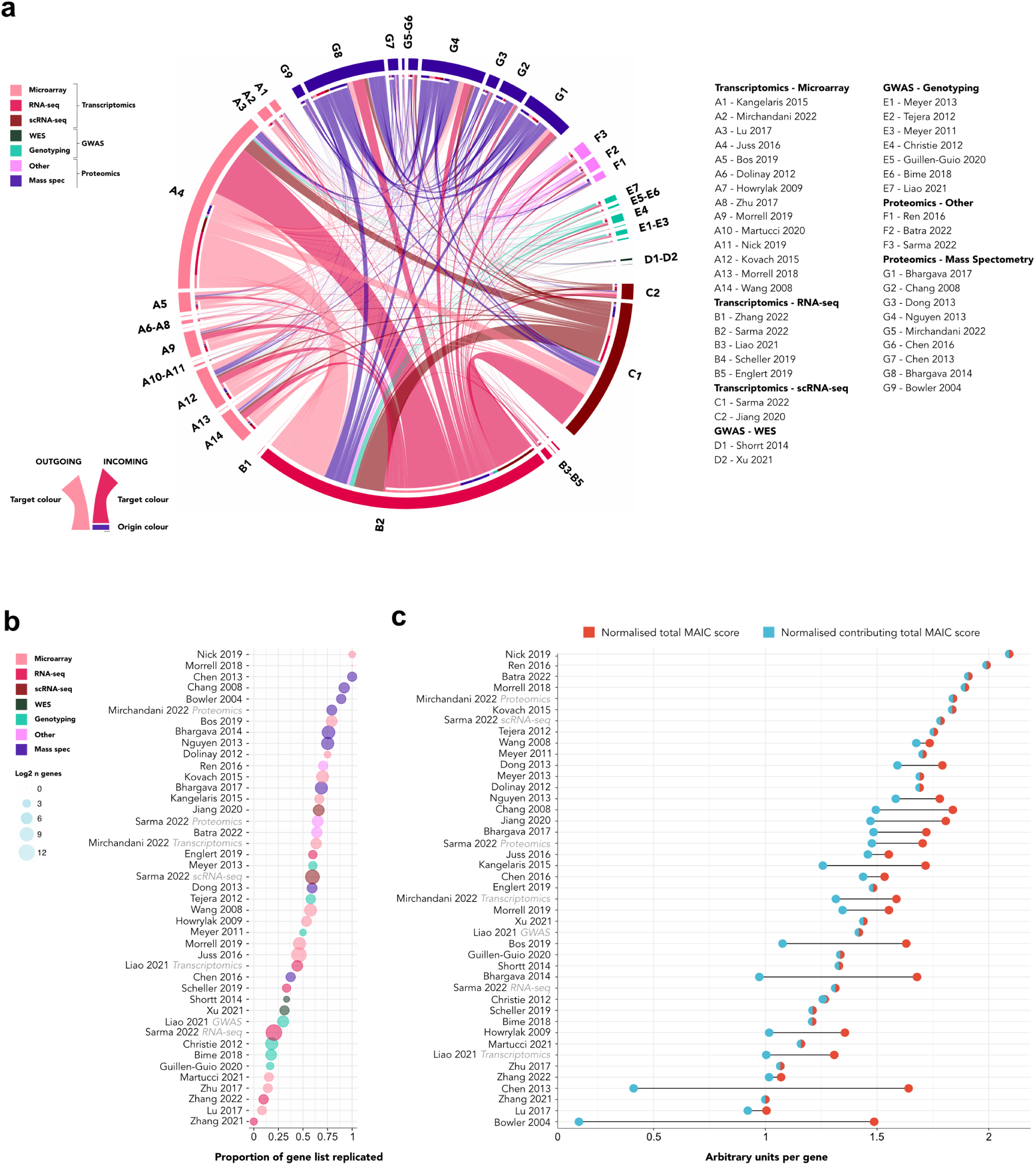
Attributing information in MAIC. (a) Shared information between gene lists. Links indicate shared summed common gene scores between studies. (b) Proportion of replicated genes. Circle diameter is equal to logarithm (base 2) of gene number per list. (c) Total MAIC score (totMS) normalised by number of genes. Overlapping circles denote equal normalised totMS and contribution (ctotMS - sum of common gene scores contributing to MAIC score for a gene), indicating all gene scores contributed to MAIC.

**Supplementary-Figure 3:**
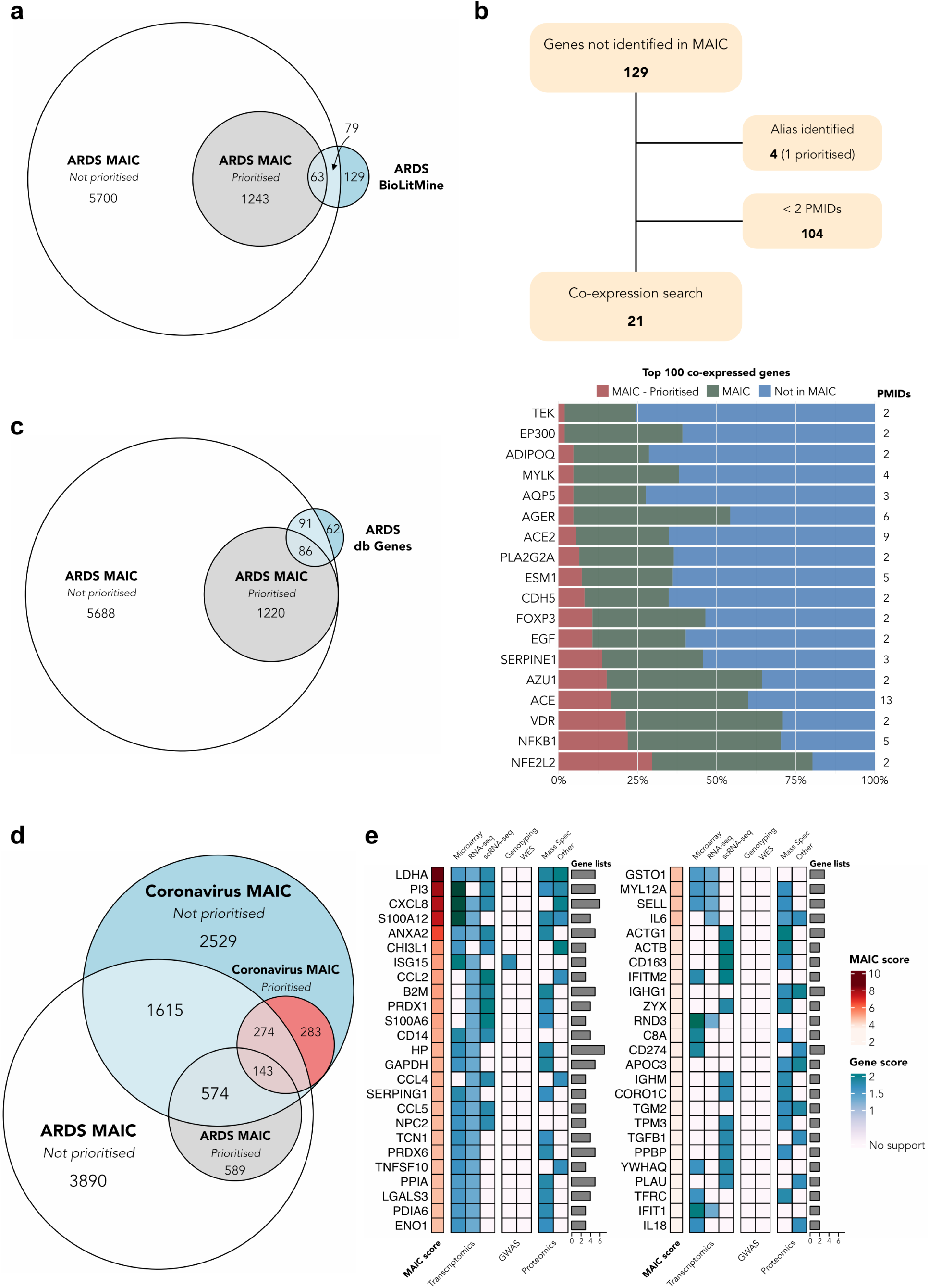
Overlap between ARDS MAIC and ARDS-associated genes and ARDS MAIC and coronavirus MAIC. (a) Euler diagram of gene overlap between ARDS MAIC and a BioLitMine search using the ARDS MeSH term. (b) Schematic overview of a co-expression search for genes identified in the BioLitMine search but not present in ARDS MAIC and a stacked bar plot of the proportion of the 100 most co-expressed genes of this group and ARDS MAIC. (c) Euler diagram of gene overlap between ARDS MAIC and the ARDS Database of Genes. (d) Euler diagram of gene overlap between ARDS MAIC and a MAIC of COVID-19 host-response studies. (e) Heatmap of the 50 top ranked ARDS MAIC genes also prioritised by the coronavirus MAIC, displaying the ARDS MAIC score for each gene, highest gene score in each category, and the number of supporting gene lists.

**Supplementary-Figure 4:**
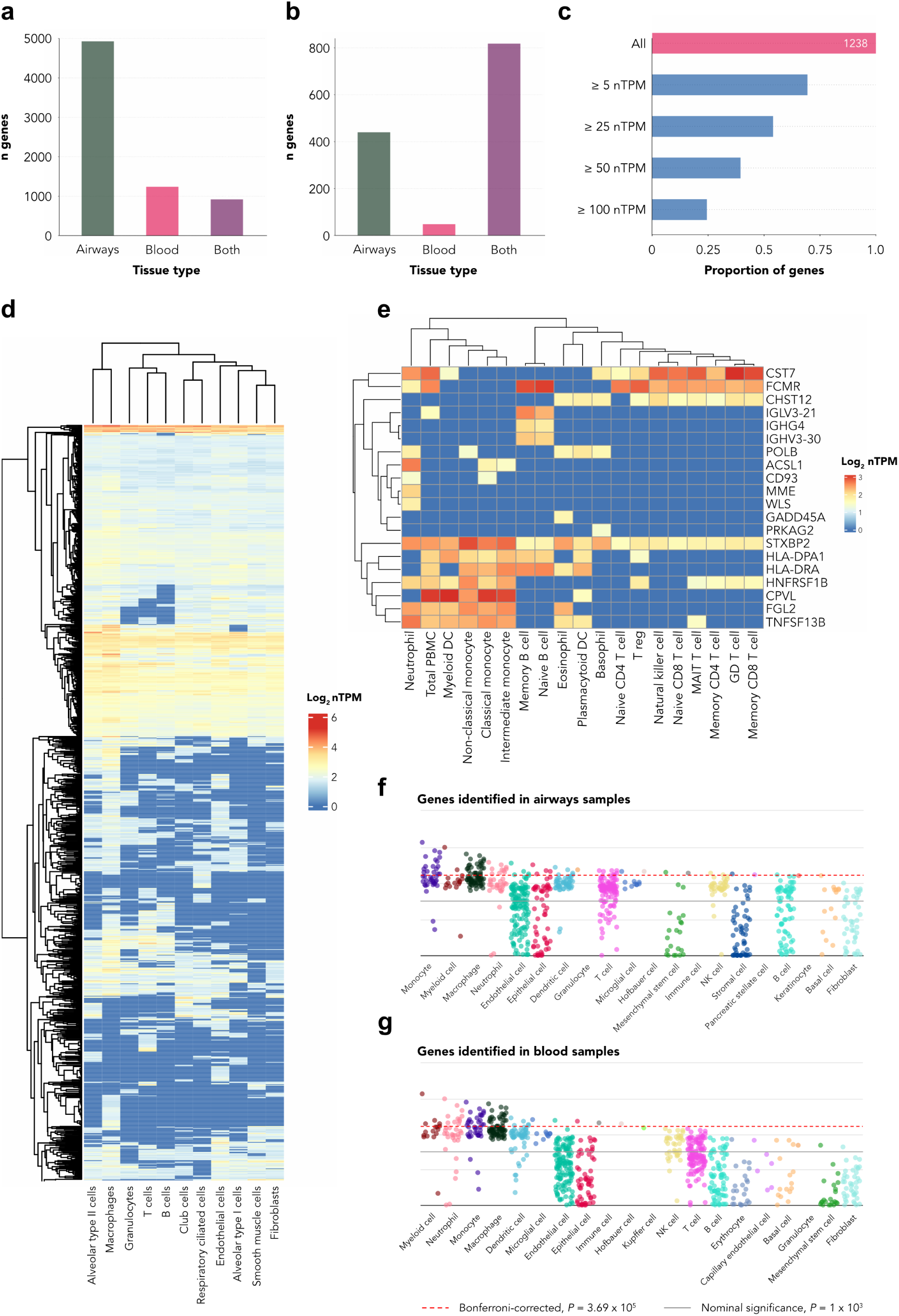
Tissue and cell-specific expression. (a) Bar plot of the tissue type in which genes are identified - all genes (n=7,085). (b) Bar plot of the tissue type in which genes are identified - prioritised genes (n=1,306). (c) Bar plot of the proportion of genes identified solely in blood meeting mRNA expression thresholds in bulk lung tissue. nTPM - normalised transcripts per million. (d) Heatmap of mRNA expression in lung cell-types for genes identified in studies based on airways sampling. (e) Heatmap of mRNA expression in blood cell-types for genes identified solely in studies based on blood sampling. (f) Manhatten plot of the top 20 cell types overenriched for expression of genes identified by studies based on airways sampling. (g) Manhatten plot of the top 20 cell types overenriched for expression of genes identified by studies based on blood sampling.

**Supplementary-Figure 5:**
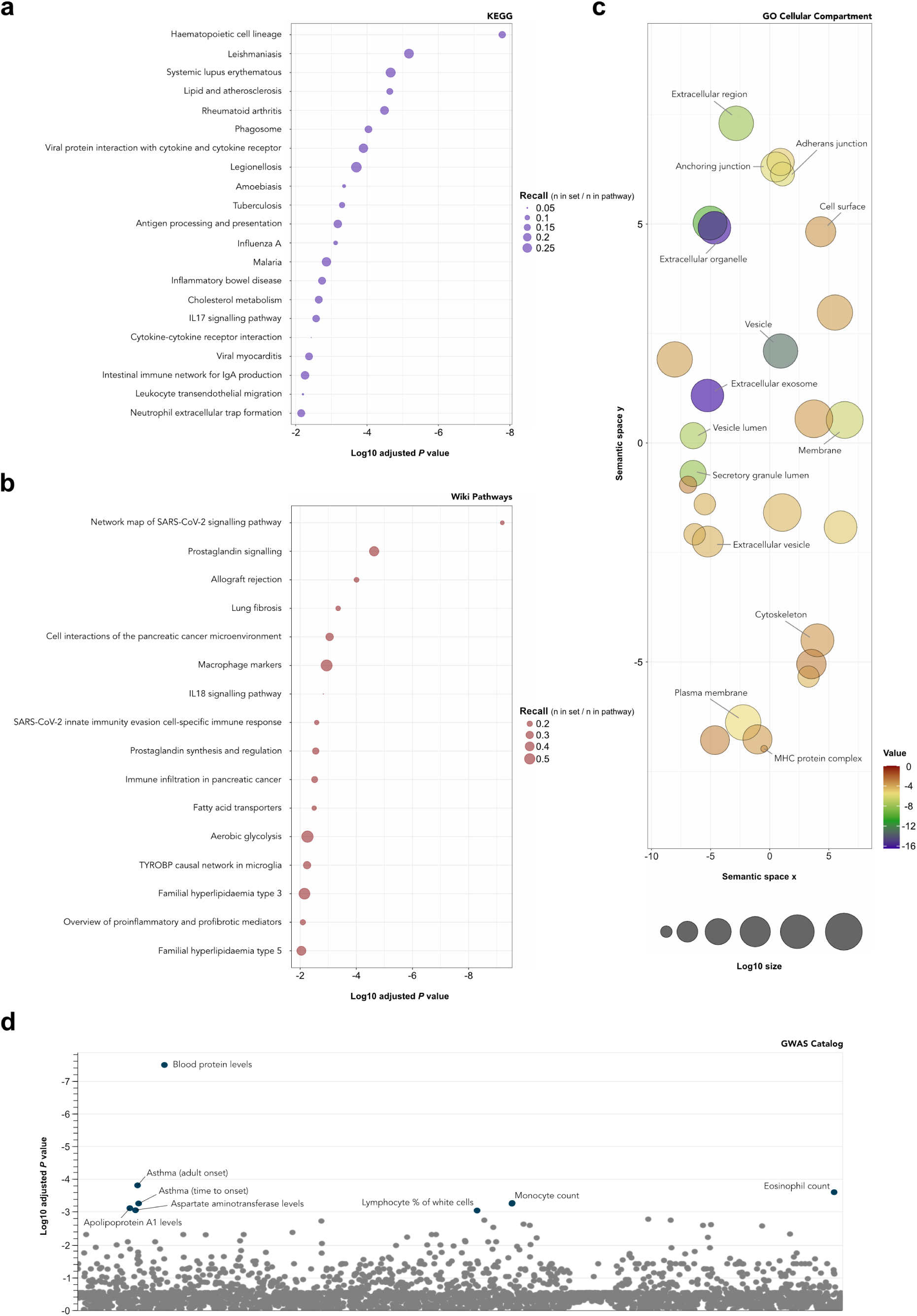
Functional enrichment. (a) Significantly enriched KEGG terms (*P* < 0.01) for prioritised genes. Terms size proportional to recall. (b) Significantly enriched WikiPathways terms (*P* < 0.01) for prioritised genes. Terms size proportional to recall. (c) Scatter plot of the semantic similarity between signficantly enriched GO cellular component terms for prioritised genes (d) Manhatten plot of the overenrichment of prioritised genes against the GWAS catolog.

**Supplementary-Figure 6:**
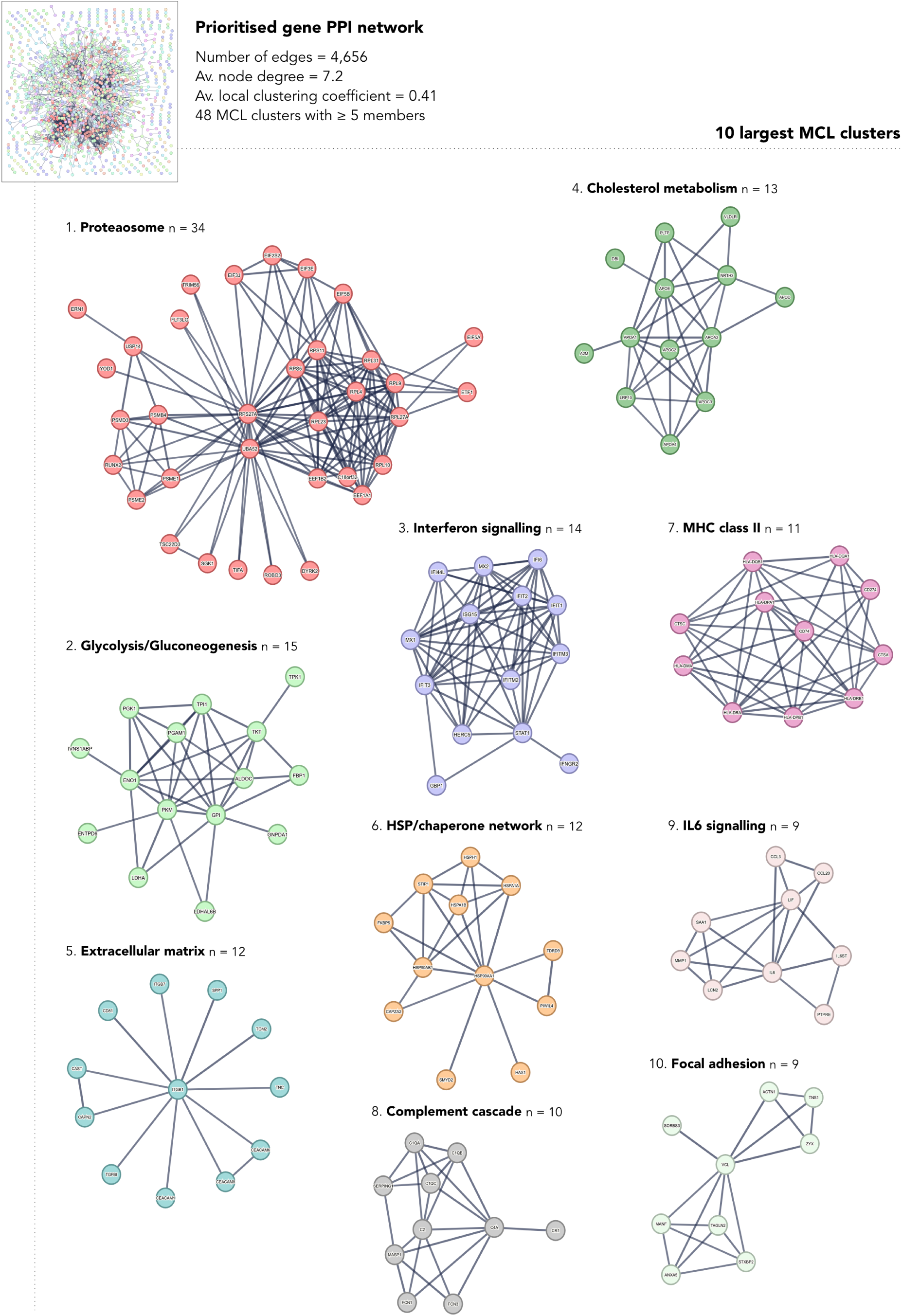
PPI clusters. A protein-protein interaction network of prioritsed genes and the 10 largest graph-based clusters. Functional annotation by hand based on a concencus of enriched Reactome, KEGG, WikiPathways, and GO Biological Process terms.

**Supplementary-Figure 7:**
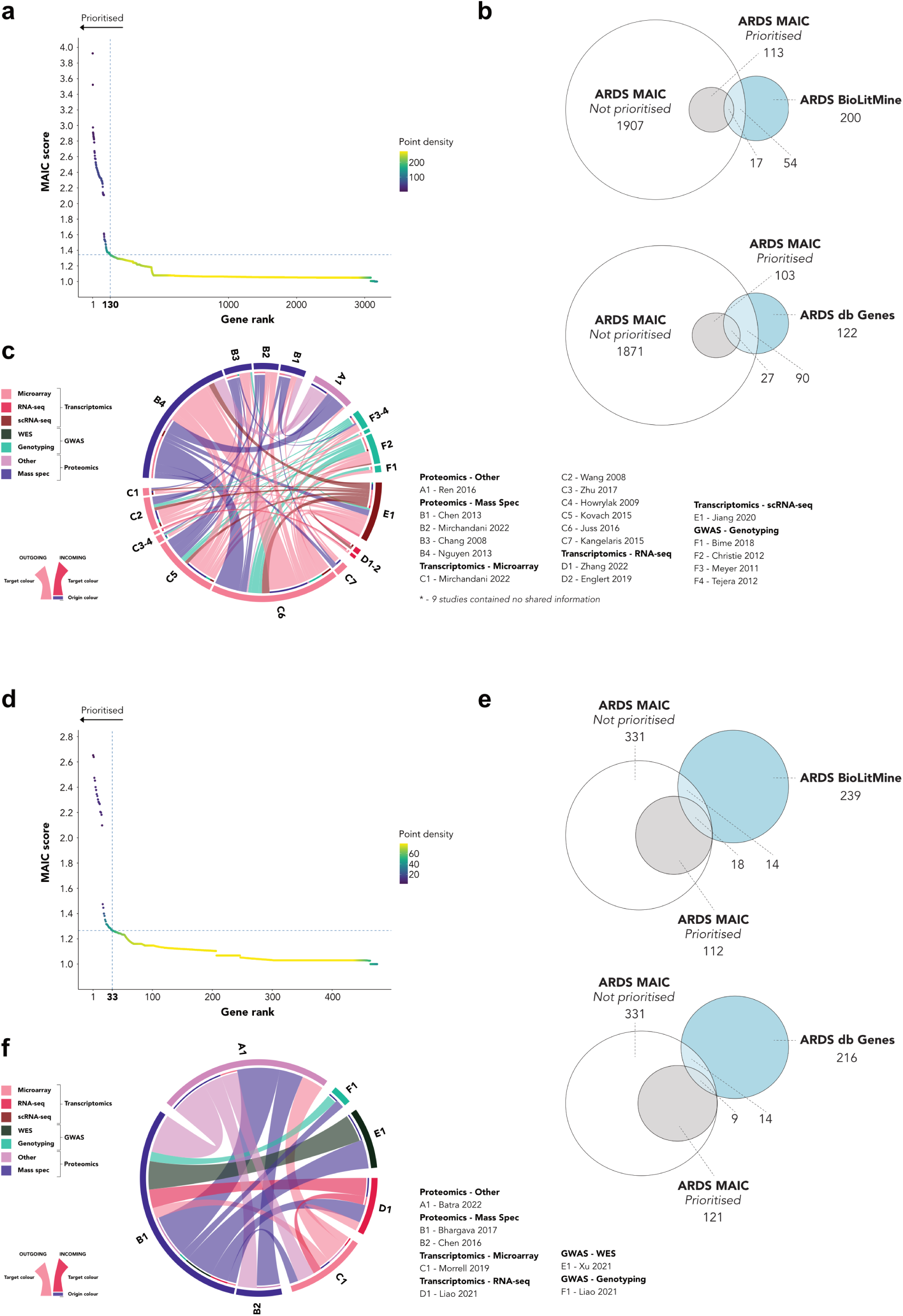
Details of MAIC on sub-groups. (a) Gene prioritisation for the ARDS MAIC ARDS vs. non-ARDS controls sub-group using the Unit Invariant Knee method. Intersection of lines identifies elbow point of best-fit curve. 130 genes in upper left quadrant were prioritied. (b) Euler diagrams of gene overlap between the ARDS vs. non-ARDS controls sub-group and a BioLitMine search using the ARDS MeSH term and the ARDS Database of Genes. (c) Shared information between ARDS vs. non-ARDS controls gene lists. Links indicate shared summed common gene scores between studies. (d) Gene prioritisation for the ARDS MAIC survival sub-group using the Unit Invariant Knee method. Intersection of lines identifies elbow point of best-fit curve. 33 genes in upper left quadrant were prioritied. (e) Euler diagrams of gene ovelap between the survival sub-group and a BioLitMine search using the ARDS MeSH term and the ARDS Database of Genes. (f) Shared information between survival gene lists. Links indicate shared

**Supplementary Table 1.** Gene list information content and contribution.

| **Study** | **Method** | **Category** | **N genes** | **totMS (% sum)** | **ctotMS (% sum)** |
| --- | --- | --- | --- | --- | --- |
| Sarma<sup>1</sup> | Transcriptomics | RNA-seq | 4954 | 50.8 | 53.1 |
| Juss<sup>2</sup> | Transcriptomics | Microarray | 1318 | 16 | 15.7 |
| Sarma<sup>1</sup> | Transcriptomics | scRNA-seq | 706 | 9.8 | 10.3 |
| Nguyen<sup>3</sup> | Proteomics | Mass Spec | 161 | 2.2 | 2.1 |
| Wang<sup>4</sup> | Transcriptomics | Microarray | 137 | 1.9 | 1.9 |
| Bhargava<sup>5</sup> | Proteomics | Mass Spec | 233 | 3.1 | 1.9 |
| Kovach<sup>6</sup> | Transcriptomics | Microarray | 123 | 1.8 | 1.9 |
| Bhargava<sup>7</sup> | Proteomics | Mass Spec | 144 | 1.9 | 1.8 |
| Morrell<sup>8</sup> | Transcriptomics | Microarray | 155 | 1.9 | 1.7 |
| Christie<sup>9</sup> | GWAS | Genotyping | 143 | 1.4 | 1.5 |
| Liao<sup>10</sup> | GWAS | Genotyping | 67 | 0.7 | 0.8 |
| Sarma<sup>1</sup> | Proteomics | Other | 60 | 0.8 | 0.7 |
| Jiang<sup>11</sup> | Transcriptomics | scRNA-seq | 53 | 0.7 | 0.6 |
| Batra<sup>12</sup> | Proteomics | Other | 39 | 0.6 | 0.6 |
| Bime<sup>13</sup> | GWAS | Genotyping | 51 | 0.5 | 0.5 |
| Bos<sup>14</sup> | Transcriptomics | Microarray | 53 | 0.7 | 0.5 |
| Chang<sup>15</sup> | Proteomics | Mass Spec | 37 | 0.5 | 0.5 |
| Mirchandani<sup>16</sup> | Transcriptomics | Microarray | 41 | 0.5 | 0.4 |
| Mirchandani<sup>16</sup> | Proteomics | Mass Spec | 29 | 0.4 | 0.4 |
| Liao<sup>10</sup> | Transcriptomics | RNA-seq | 43 | 0.4 | 0.4 |
| Dong<sup>17</sup> | Proteomics | Mass Spec | 27 | 0.4 | 0.4 |
| Ren<sup>18</sup> | Proteomics | Other | 17 | 0.3 | 0.3 |
| Tejera<sup>19</sup> | GWAS | Genotyping | 19 | 0.3 | 0.3 |
| Howrylak<sup>20</sup> | Transcriptomics | Microarray | 28 | 0.3 | 0.2 |
| Xu<sup>21</sup> | GWAS | WES | 16 | 0.2 | 0.2 |
| Chen<sup>22</sup> | Proteomics | Mass Spec | 16 | 0.2 | 0.2 |
| Zhang<sup>23</sup> | Transcriptomics | RNA-seq | 20 | 0.2 | 0.2 |
| Kangelaris<sup>24</sup> | Transcriptomics | Microarray | 15 | 0.2 | 0.2 |
| Meyer<sup>25</sup> | GWAS | Genotyping | 10 | 0.1 | 0.1 |
| Martucci<sup>26</sup> | Transcriptomics | Microarray | 13 | 0.1 | 0.1 |
| Zhu<sup>27</sup> | Transcriptomics | Microarray | 14 | 0.1 | 0.1 |
| Englert<sup>28</sup> | Transcriptomics | RNA-seq | 10 | 0.1 | 0.1 |
| Lu<sup>29</sup> | Transcriptomics | Microarray | 12 | < 0.1 | < 0.1 |
| Scheller<sup>30</sup> | Transcriptomics | RNA-seq | 9 | < 0.1 | < 0.1 |
| Nick<sup>31</sup> | Transcriptomics | Microarray | 4 | < 0.1 | < 0.1 |
| Guillen-Guio<sup>32</sup> | GWAS | Genotyping | 6 | < 0.1 | < 0.1 |
| Meyer<sup>33</sup> | GWAS | Genotyping | 4 | < 0.1 | < 0.1 |
| Dolinay<sup>34</sup> | Transcriptomics | Microarray | 4 | < 0.1 | < 0.1 |
| Chen<sup>35</sup> | Proteomics | Mass Spec | 16 | < 0.1 | < 0.1 |
| Zhang<sup>36</sup> | Transcriptomics | RNA-seq | 5 | < 0.1 | < 0.1 |
| Shortt<sup>37</sup> | GWAS | WES | 3 | < 0.1 | < 0.1 |
| Bowler<sup>38</sup> | Proteomics | Mass Spec | 18 | < 0.1 | < 0.1 |
| Morrell<sup>39</sup> | Transcriptomics | Microarray | 1 | < 0.1 | < 0.1 |

Abbreviations: GWAS - Genome-wide association study; Mass Spec - Mass spectrometry; totMS - Total MAIC score; ctotMS - Contributing total MAIC score; WES - Whole-exome sequencing. totMS and ctotMS are reported as the percentage of the sum of totMS/ctotMS for all lists included in the analysis.

**Supplementary Table 2.** ARDS MAIC prioritised genes found in common by BioLitMine with >= 2 associated publications.

| Gene | Publication |  | MAIC rank |
| --- | --- | --- | --- |
|  | count | PubMed IDs |  |
| TGFB1 | 8 | 30395619, 29083412, 28188225, 27309347, 22034170, 20142324, 16100012, 12654639 | 225 |
| VEGFA | 8 | 24356493, 23542734, 21797753, 19543148, 19349383, 17289863, 15920019, 15741444 | 320 |
| IL10 | 8 | 32217834, 31936183, 30280795, 28432351, 22033829, 21138342, 18242340, 16585075 | 1268 |
| SFTPB | 6 | 21128671, 18679120, 16100012, 15190959, 14718442, 12490037 | 177 |
| IL17A | 6 | 34239039, 32795834, 32651218, 30655311, 26709006, 26002979 | 1294 |
| PI3 | 5 | 28187039, 24617927, 19251943, 19197381, 18203972 | 2 |
| CXCL8 | 5 | 22897124, 22080750, 21348591, 17498967, 14729508 | 3 |
| IL6 | 5 | 34757857, 33250487, 32826331, 31261506, 18593632 | 144 |
| TNF | 5 | 31261506, 22507624, 21784970, 17034639, 16135717 | 651 |
| NAMPT | 4 | 24821571, 24053186, 18486613, 17392604 | 58 |
| IL1RN | 4 | 30095747, 29943912, 23449693, 18838927 | 175 |
| SCGB1A1 | 4 | 32787812, 28548310, 18521628, 16215398 | 187 |
| NPPB | 4 | 28322314, 26359292, 21696613, 19830720 | 1239 |
| HGF | 3 | 18065658, 17702746, 11943656 | 343 |
| IL33 | 3 | 33936076, 31147742, 23000728 | 385 |
| CXCL10 | 3 | 31651197, 23542734, 23144331 | 671 |
| S100A12 | 2 | 26274928, 24887223 | 5 |
| MUC1 | 2 | 21418654, 17565019 | 69 |
| PLAU | 2 | 23064953, 17994220 | 244 |
| EPAS1 | 2 | 28613249, 25574837 | 425 |
| FASLG | 2 | 30385692, 12414525 | 503 |
| EDN1 | 2 | 27765761, 17875064 | 643 |
| AKT1 | 2 | 27607575, 15961723 | 950 |
| MMP8 | 2 | 24651234, 15187163 | 1223 |

**Supplementary Table 3.** ARDS susceptibility gene list information content and contribution.

| **Study** | **Method** | **Category** | **N genes** | **totMS (% sum)** | **ctotMS (% sum)** |
| --- | --- | --- | --- | --- | --- |
| Juss<sup>2</sup> | Transcriptomics | Microarray | 1318 | 54.7 | 54.7 |
| Nguyen<sup>3</sup> | Proteomics | Mass Spec | 161 | 8.1 | 7.7 |
| Christie<sup>9</sup> | GWAS | Genotyping | 143 | 6 | 6.3 |
| Kovach<sup>6</sup> | Transcriptomics | Microarray | 123 | 5.8 | 6.1 |
| Wang<sup>4</sup> | Transcriptomics | Microarray | 137 | 5.8 | 6 |
| Jiang<sup>11</sup> | Transcriptomics | scRNA-seq | 53 | 2.9 | 3 |
| Bime<sup>13</sup> | GWAS | Genotyping | 51 | 2.2 | 2.3 |
| Mirchandani<sup>16</sup> | Transcriptomics | Microarray | 41 | 1.7 | 1.6 |
| Chang<sup>15</sup> | Proteomics | Mass Spec | 37 | 1.9 | 1.5 |
| Mirchandani<sup>16</sup> | Proteomics | Mass Spec | 29 | 1.4 | 1.3 |
| Howrylak<sup>20</sup> | Transcriptomics | Microarray | 28 | 1.2 | 1.3 |
| Ren<sup>18</sup> | Proteomics | Other | 17 | 1 | 1.1 |
| Tejera<sup>19</sup> | GWAS | Genotyping | 19 | 0.9 | 1 |
| Chen<sup>35</sup> | Proteomics | Mass Spec | 16 | 0.9 | 0.9 |
| Zhang<sup>23</sup> | Transcriptomics | RNA-seq | 20 | 0.8 | 0.9 |
| Zhu<sup>27</sup> | Transcriptomics | Microarray | 14 | 0.6 | 0.6 |
| Kangelaris<sup>24</sup> | Transcriptomics | Microarray | 15 | 0.7 | 0.6 |
| Englert<sup>28</sup> | Transcriptomics | RNA-seq | 10 | 0.6 | 0.6 |
| Lu<sup>29</sup> | Transcriptomics | Microarray | 12 | 0.5 | 0.5 |
| Meyer<sup>25</sup> | GWAS | Genotyping | 10 | 0.4 | 0.4 |
| Bowler<sup>38</sup> | Proteomics | Mass Spec | 18 | 0.9 | 0.4 |
| Scheller<sup>30</sup> | Transcriptomics | RNA-seq | 9 | 0.4 | 0.3 |
| Guillen-Guio<sup>32</sup> | GWAS | Genotyping | 6 | 0.2 | 0.2 |
| Zhang<sup>36</sup> | Transcriptomics | RNA-seq | 5 | 0.2 | 0.2 |
| Dolinay<sup>34</sup> | Transcriptomics | Microarray | 4 | 0.2 | 0.2 |
| Shortt<sup>37</sup> | GWAS | WES | 3 | 0.1 | 0.1 |
| Meyer<sup>33</sup> | GWAS | Genotyping | 4 | < 0.1 | 0.1 |
| Morrell<sup>39</sup> | Transcriptomics | Microarray | 1 | < 0.1 | < 0.1 |

Abbreviations: GWAS - Genome-wide association study; Mass Spec - Mass spectrometry; totMS - Total MAIC score; ctotMS - Contributing total MAIC score; WES - Whole-exome sequencing. totMS and ctotMS are reported as the percentage of the sum of totMS/ctotMS for all lists included in the analysis.

**Supplementary Table 4.** ARDS survival/severity gene list information content and contribution.

| **Study** | **Method** | **Category** | **N genes** | **totMS (% sum)** | **ctotMS (% sum)** |
| --- | --- | --- | --- | --- | --- |
| Bhargava<sup>7</sup> | Proteomics | Mass Spec | 144 | 30.4 | 30.3 |
| Morrell<sup>8</sup> | Transcriptomics | Microarray | 155 | 29.7 | 29.7 |
| Liao<sup>10</sup> | GWAS | Genotyping | 67 | 12.9 | 13 |
| Batra<sup>12</sup> | Proteomics | Other | 39 | 9.4 | 9.4 |
| Liao<sup>10</sup> | Transcriptomics | RNA-seq | 43 | 8.5 | 8.5 |
| Xu<sup>21</sup> | GWAS | WES | 16 | 3.5 | 3.5 |
| Chen<sup>22</sup> | Proteomics | Mass Spec | 16 | 3.4 | 3.4 |
| Lu<sup>29</sup> | Transcriptomics | Microarray | 12 | 2.2 | 2.2 |

Abbreviations: GWAS - Genome-wide association study; Mass Spec - Mass spectrometry; totMS - Total MAIC score; ctotMS - Contributing total MAIC score; WES - Whole-exome sequencing. totMS and ctotMS are reported as the percentage of the sum of totMS/ctotMS for all lists included in the analysis.

### Supplementary Data

**Supplementary Data File 1. Raw gene list input to MAIC**. https://github.com/JonathanEMillar/ards_maic_manuscript/Supplementary_Data_File_1.csv

**Supplementary Data File 2. MAIC output - overall**. https://github.com/JonathanEMillar/ards_maic_manuscript/Supplementary_Data_File_2.csv

**Supplementary Data File 3. BioLitMine and ARDS Database of Genes results**. https://github.com/JonathanEMillar/ards_maic_manuscript/Supplementary_Data_File_3.csv

**Supplementary Data File 4. MAIC output - ARDS vs. non-ARDS controls sub-group**. https://github.com/JonathanEMillar/ards_maic_manuscript/Supplementary_Data_File_4.csv

**Supplementary Data File 5. MAIC output - survival sub-group**. https://github.com/JonathanEMillar/ards_maic_manuscript/Supplementary_Data_File_5.csv

**Supplementary Data File 6. Functional enrichment results - overall**. https://github.com/JonathanEMillar/ards_maic_manuscript/Supplementary_Data_File_6.csv

**Supplementary Data File 7. Functional enrichment results - ARDS vs. non-ARDS controls sub-group**. https://github.com/JonathanEMillar/ards_maic_manuscript/Supplementary_Data_File_7.csv

**Supplementary Data File 8. Functional enrichment results - survival sub-group**. https://github.com/JonathanEMillar/ards_maic_manuscript/Supplementary_Data_File_8.csv

## Glossary

MAIC score: the score assigned by MAIC to a given gene considering all lists.
Gene score: the score assigned by MAIC to a given gene in a given list.
Total MAIC score: the sum of all scores assigned by MAIC to a genes in a given list.
Contributing total MAIC score: the sum of all scores assigned by MAIC to a genes in a given list where that score contributes to the MAIC score for that gene (i.e., excluding those gene scores that are not used because a gene score from another list in the same category is greater).

## References

1. ARDS Definition Task Force et al. Acute respiratory distress syndrome: The berlin definition. JAMA 307, 2526–2533 (2012).

2. Bellani, G. et al. Epidemiology, patterns of care, and mortality for patients with acute respiratory distress syndrome in intensive care units in 50 countries. JAMA 315, 788–800 (2016).

3. Wilson, J. G. & Calfee, C. S. ARDS subphenotypes: Understanding a heterogeneous syndrome. Crit. Care 24, 102 (2020).

4. Laffey, J. G. & Kavanagh, B. P. Negative trials in critical care: Why most research is probably wrong. Lancet Respir. Med. 6, 659–660 (2018).

5. Bos, L. D. J. et al. Towards a biological definition of ARDS: Are treatable traits the solution? Intensive Care Med. Exp. 10, 8 (2022).

6. Peter W Horby, and et al. Baricitinib in patients admitted to hospital with COVID-19 (RECOVERY): A randomised, controlled, open-label, platform trial and updated meta-analysis. (2022) doi:10.1101/2022.03.02.22271623.

7. Richardson, P. et al. Baricitinib as potential treatment for 2019-nCoV acute respiratory disease. Lancet 395, e30–e31 (2020).

8. Pairo-Castineira, E. et al. Genetic mechanisms of critical illness in COVID-19. Nature 591, 92–98 (2021).

9. Gomez-Cabrero, D. et al. Data integration in the era of omics: Current and future challenges. BMC Syst. Biol. 8 Suppl 2, I1 (2014).

10. Li, B. et al. Genome-wide CRISPR screen identifies host dependency factors for influenza a virus infection. Nat. Commun. 11, 164 (2020).

11. Parkinson, N. et al. Dynamic data-driven meta-analysis for prioritisation of host genes implicated in COVID-19. Sci. Rep. 10, 22303 (2020).

12. Wang, B. et al. Systematic comparison of ranking aggregation methods for gene lists in experimental results. Bioinformatics 38, 4927–4933 (2022).

13. Batra, R. et al. Multi-omic comparative analysis of COVID-19 and bacterial sepsis-induced ARDS. PLoS Pathog. 18, e1010819 (2022).

14. Bhargava, M. et al. Proteomic profiles in acute respiratory distress syndrome differentiates survivors from non-survivors. PLoS One 9, e109713 (2014).

15. Bhargava, M. et al. Bronchoalveolar lavage fluid protein expression in acute respiratory distress syndrome provides insights into pathways activated in subjects with different outcomes. Sci. Rep. 7, 7464 (2017).

16. Bime, C. et al. Genome-wide association study in african americans with acute respiratory distress syndrome identifies the selectin P ligand gene as a risk factor. Am. J. Respir. Crit. Care Med. 197, 1421–1432 (2018).

17. Bos, L. D. J. et al. Understanding heterogeneity in biologic phenotypes of acute respiratory distress syndrome by leukocyte expression profiles. Am. J. Respir. Crit. Care Med. 200, 42–50 (2019).

18. Bowler, R. P. et al. Proteomic analysis of pulmonary edema fluid and plasma in patients with acute lung injury. Am. J. Physiol. Lung Cell. Mol. Physiol. 286, L1095–104 (2004).

19. Chang, D. W. et al. Proteomic and computational analysis of bronchoalveolar proteins during the course of the acute respiratory distress syndrome. Am. J. Respir. Crit. Care Med. 178, 701–709 (2008).

20. Chen, X., Shan, Q., Jiang, L., Zhu, B. & Xi, X. Quantitative proteomic analysis by iTRAQ for identification of candidate biomarkers in plasma from acute respiratory distress syndrome patients. Biochem. Biophys. Res. Commun. 441, 1–6 (2013).

21. Chen, C., Shi, L., Li, Y., Wang, X. & Yang, S. Disease-specific dynamic biomarkers selected by integrating inflammatory mediators with clinical informatics in ARDS patients with severe pneumonia. Cell Biol. Toxicol. 32, 169–184 (2016).

22. Christie, J. D. et al. Genome wide association identifies PPFIA1 as a candidate gene for acute lung injury risk following major trauma. PLoS One 7, e28268 (2012).

23. Dolinay, T. et al. Inflammasome-regulated cytokines are critical mediators of acute lung injury. Am. J. Respir. Crit. Care Med. 185, 1225–1234 (2012).

24. Dong, H. et al. Comparative analysis of the alveolar macrophage proteome in ALI/ARDS patients between the exudative phase and recovery phase. BMC Immunol. 14, 25 (2013).

25. Englert, J. A. et al. Whole blood RNA sequencing reveals a unique transcriptomic profile in patients with ARDS following hematopoietic stem cell transplantation. Respir. Res. 20, 15 (2019).

26. Frenzel, J. et al. Outcome prediction in pneumonia induced ALI/ARDS by clinical features and peptide patterns of BALF determined by mass spectrometry. PLoS One 6, e25544 (2011).

27. Guillen-Guio, B. et al. Sepsis-associated acute respiratory distress syndrome in individuals of european ancestry: A genome-wide association study. Lancet Respir. Med. 8, 258–266 (2020).

28. Howrylak, J. A. et al. Discovery of the gene signature for acute lung injury in patients with sepsis. Physiol. Genomics 37, 133–139 (2009).

29. Jiang, Y., et al. Single cell RNA sequencing identifies an early monocyte gene signature in acute respiratory distress syndrome. JCI Insight 5, (2020).

30. Juss, J. K. et al. Acute respiratory distress syndrome neutrophils have a distinct phenotype and are resistant to phosphoinositide 3-kinase inhibition. Am. J. Respir. Crit. Care Med. 194, 961–973 (2016).

31. Kangelaris, K. N. et al. Increased expression of neutrophil-related genes in patients with early sepsis-induced ARDS. Am. J. Physiol. Lung Cell. Mol. Physiol. 308, L1102–13 (2015).

32. Kovach, M. A. et al. Microarray analysis identifies IL-1 receptor type 2 as a novel candidate biomarker in patients with acute respiratory distress syndrome. Respir. Res. 16, 29 (2015).

33. Liao, S. Y. et al. Identification of early and intermediate biomarkers for ARDS mortality by multi-omic approaches. Sci. Rep. 11, 18874 (2021).

34. Lu, X.-G. et al. Circulating miRNAs as biomarkers for severe acute pancreatitis associated with acute lung injury. World J. Gastroenterol. 23, 7440–7449 (2017).

35. Martucci, G. et al. Identification of a circulating miRNA signature to stratify acute respiratory distress syndrome patients. J. Pers. Med. 11, 15 (2020).

36. Meyer, N. J. et al. ANGPT2 genetic variant is associated with trauma-associated acute lung injury and altered plasma angiopoietin-2 isoform ratio. Am. J. Respir. Crit. Care Med. 183, 1344–1353 (2011).

37. Meyer, N. J. et al. IL1RN coding variant is associated with lower risk of acute respiratory distress syndrome and increased plasma IL-1 receptor antagonist. Am. J. Respir. Crit. Care Med. 187, 950–959 (2013).

38. Mirchandani, A. S. et al. Hypoxia shapes the immune landscape in lung injury and promotes the persistence of inflammation. Nat. Immunol. 23, 927–939 (2022).

39. Morrell, E. D. et al. Cytometry TOF identifies alveolar macrophage subtypes in acute respiratory distress syndrome. JCI Insight 3, (2018).

40. Morrell, E. D. et al. Alveolar macrophage transcriptional programs are associated with outcomes in acute respiratory distress syndrome. Am. J. Respir. Crit. Care Med. 200, 732–741 (2019).

41. Nick, J. A. et al. Extremes of interferon-stimulated gene expression associate with worse outcomes in the acute respiratory distress syndrome. PLoS One 11, e0162490 (2016).

42. Nguyen, E. V. et al. Proteomic profiling of bronchoalveolar lavage fluid in critically ill patients with ventilator-associated pneumonia. PLoS One 8, e58782 (2013).

43. Ren, S. et al. Deleted in malignant brain tumors 1 protein is a potential biomarker of acute respiratory distress syndrome induced by pneumonia. Biochem. Biophys. Res. Commun. 478, 1344–1349 (2016).

44. Sarma, A., et al. Hyperinflammatory ARDS is characterized by interferon-stimulated gene expression, t-cell activation, and an altered metatranscriptome in tracheal aspirates. bioRxiv (2022).

45. Scheller, N. et al. Proviral MicroRNAs detected in extracellular vesicles from bronchoalveolar lavage fluid of patients with influenza virus-induced acute respiratory distress syndrome. J. Infect. Dis. 219, 540–543 (2019).

46. Shortt, K. et al. Identification of novel single nucleotide polymorphisms associated with acute respiratory distress syndrome by exome-seq. PLoS One 9, e111953 (2014).

47. Wang, Z., Beach, D., Su, L., Zhai, R. & Christiani, D. C. A genome-wide expression analysis in blood identifies pre-elafin as a biomarker in ARDS. Am. J. Respir. Cell Mol. Biol. 38, 724–732 (2008).

48. Tejera, P. et al. Distinct and replicable genetic risk factors for acute respiratory distress syndrome of pulmonary or extrapulmonary origin. J. Med. Genet. 49, 671–680 (2012).

49. Xu, J.-Y. et al. Nucleotide polymorphism in ARDS outcome: A whole exome sequencing association study. Ann. Transl. Med. 9, 780 (2021).

50. Zhang, S. et al. miR-584 and miR-146 are candidate biomarkers for acute respiratory distress syndrome. Exp. Ther. Med. 21, 445 (2021).

51. Zhang, C. et al. Differential expression profile of plasma exosomal microRNAs in acute type a aortic dissection with acute lung injury. Sci. Rep. 12, 11667 (2022).

52. Zhu, Z. et al. Whole blood microRNA markers are associated with acute respiratory distress syndrome. Intensive Care Med. Exp. 5, 38 (2017).

53. Christopoulos, D. Introducing unit invariant knee (UIK) as an objective choice for elbow point in multivariate data analysis techniques. SSRN Electron. J. (2016).

54. Hu, Y. et al. BioLitMine: Advanced mining of biomedical and biological literature about human genes and genes from major model organisms. G3 (Bethesda) 10, 4531–4539 (2020).

55. Quintanilla, E., Diwa, K., Nguyen, A., Vu, L. & Toby, I. T. A data report on the curation and development of a database of genes for acute respiratory distress syndrome. Front. Genet. 12, 750568 (2021).

56. Lachmann, A. et al. Massive mining of publicly available RNA-seq data from human and mouse. Nat. Commun. 9, 1366 (2018).

57. Fischer, M. & Hoffmann, S. Synthesizing genome regulation data with vote-counting. Trends Genet 38, 1208– 1216 (2022).

58. Bos, L. D. J. et al. Towards a biological definition of ARDS: are treatable traits the solution? Intensive Care Med Exp 10, 8 (2022).

59. Kolde, R., Laur, S., Adler, P. & Vilo, J. Robust rank aggregation for gene list integration and meta-analysis. Bioinformatics 28, 573–580 (2012).

60. Bachofen, M. & Weibel, E. R. Alterations of the gas exchange apparatus in adult respiratory insufficiency associated with septicemia. Am Rev Respir Dis 116, 589–615 (1977).

61. Cambier, S., Gouwy, M. & Proost, P. The chemokines CXCL8 and CXCL12: molecular and functional properties, role in disease and efforts towards pharmacological intervention. Cell Mol Immunol 20, 217–251 (2023).

62. Moore, A. R. et al. Elevated Plasma Interleukin-18 Identifies High-Risk Acute Respiratory Distress Syndrome Patients not Distinguished by Prior Latent Class Analyses Using Traditional Inflammatory Cytokines: A Retrospective Analysis of Two Randomized Clinical Trials. Crit Care Med (2023).

63. Davey, A., McAuley, D. F. & O’Kane, C. M. Matrix metalloproteinases in acute lung injury: mediators of injury and drivers of repair. Eur Respir J 38, 959–970 (2011).

64. Ballester, B., Milara, J. & Cortijo, J. The role of mucin 1 in respiratory diseases. Eur Respir Rev 30, (2021).

65. Weiske, J. & Huber, O. The histidine triad protein Hint1 triggers apoptosis independent of its enzymatic activity. J Biol Chem 281, 27356–27366 (2006).

66. Wang, X., Zhou, M. & Jiang, L. The oncogenic and immunological roles of histidine triad nucleotide-binding protein 1 in human cancers and their experimental validation in the MCF-7 cell line. Ann Transl Med 11, 147 (2023).

67. Galazka, G. et al. HINT1 peptide/Hsp70 complex induces NK-cell-dependent immunoregulation in a model of autoimmune demyelination. Eur J Immunol 44, 3026–3044 (2014).

68. Gowdy, K. M. & Fessler, M. B. Emerging roles for cholesterol and lipoproteins in lung disease. Pulm Pharmacol Ther 26, 430–437 (2013).

69. Hofmaenner, D. A., Kleyman, A., Press, A., Bauer, M. & Singer, M. The Many Roles of Cholesterol in Sepsis: A Review. Am J Respir Crit Care Med 205, 388–396 (2022).

70. Pienkos, S. M. et al. Effect of total cholesterol and statin therapy on mortality in ARDS patients: a secondary analysis of the SAILS and HARP-2 trials. Crit Care 27, 126 (2023).

71. Metkus, T. S. et al. Plasma Proprotein Convertase Subtilisin/kexin Type 9 (PCSK9) in the Acute Respiratory Distress Syndrome. Front Med (Lausanne) 9, 876046 (2022).

72. Hadjadj, J. et al. Impaired type I interferon activity and inflammatory responses in severe COVID-19 patients. Science 369, 718–724 (2020).

73. Matzaraki, V., Kumar, V., Wijmenga, C. & Zhernakova, A. The MHC locus and genetic susceptibility to autoimmune and infectious diseases. Genome Biol 18, 76 (2017).

74. ller, A. M., Cronen, C., ller, K. M. & Kirkpatrick, C. J. Heterogeneous expression of cell adhesion molecules by endothelial cells in ARDS. J Pathol 198, 270–275 (2002).

75. Demaria, O. et al. Identification of druggable inhibitory immune checkpoints on Natural Killer cells in COVID-19. Cell Mol Immunol 17, 995–997 (2020).

76. Menon, D. K. & Rosand, J. Finding a Place for Candidate Gene Studies in a Genome-Wide Association Study World. JAMA Netw Open 4, e2118594 (2021).

77. Sarma, A. et al. Tracheal aspirate RNA sequencing identifies distinct immunological features of COVID-19 ARDS. Nat Commun 12, 5152 (2021).

78. Page, M. J. et al. The PRISMA 2020 statement: An updated guideline for reporting systematic reviews. BMJ 372, n71 (2021).

79. Clark, J. et al. A full systematic review was completed in 2 weeks using automation tools: A case study. J. Clin. Epidemiol. 121, 81–90 (2020).

80. Battaglini, D. et al. Personalized medicine using omics approaches in acute respiratory distress syndrome to identify biological phenotypes. Respir. Res. 23, 318 (2022).

81. Hernández-Beeftink, T., Guillen-Guio, B., Villar, J. & Flores, C. Genomics and the acute respiratory distress syndrome: Current and future directions. Int. J. Mol. Sci. 20, 4004 (2019).

82. Reilly, J. P., Christie, J. D. & Meyer, N. J. Fifty years of research in ARDS. Genomic contributions and opportunities. Am. J. Respir. Crit. Care Med. 196, 1113–1121 (2017).

83. Zheng, F. et al. Novel biomarkers for acute respiratory distress syndrome: Genetics, epigenetics and transcriptomics. Biomark. Med. 16, 217–231 (2022).

84. Wang, B. et al. Systematic comparison of ranking aggregation methods for gene lists in experimental results. bioRxiv (2022).

85. Christopoulos, D. T. Inflection: Finds the Inflection Point of a Curve. (2019).

86. Uhlen, M. et al. Towards a knowledge-based human protein atlas. Nat. Biotechnol. 28, 1248–1250 (2010).

87. Uhlen, M. et al. A genome-wide transcriptomic analysis of protein-coding genes in human blood cells. Science 366, eaax9198 (2019).

88. Uhlén, M. et al. Proteomics. Tissue-based map of the human proteome. Science 347, 1260419 (2015).

89. Dai, Y. et al. WebCSEA: web-based cell-type-specific enrichment analysis of genes. Nucleic Acids Research 50, W782–W790 (2022).

90. Raudvere, U. et al. G:profiler: A web server for functional enrichment analysis and conversions of gene lists (2019 update). Nucleic Acids Res. 47, W191–W198 (2019).

91. Kanehisa, M. & Goto, S. KEGG: Kyoto encyclopedia of genes and genomes. Nucleic Acids Res. 28, 27–30 (2000).

92. Gillespie, M. et al. The reactome pathway knowledgebase 2022. Nucleic Acids Res. 50, D687–D692 (2022).

93. Martens, M. et al. WikiPathways: Connecting communities. Nucleic Acids Res. 49, D613–D621 (2021).

94. Ashburner, M. et al. Gene ontology: tool for the unification of biology. The Gene Ontology Consortium. Nat Genet 25, 25–29 (2000).

95. Supek, F., njak, M., kunca, N. & muc, T. REVIGO summarizes and visualizes long lists of gene ontology terms. PLoS One 6, e21800 (2011).

96. Welter, D. et al. The NHGRI GWAS Catalog, a curated resource of SNP-trait associations. Nucleic Acids Research 42, D1001–D1006 (2013).

97. Kuleshov, M. V. et al. Enrichr: a comprehensive gene set enrichment analysis web server 2016 update. Nucleic Acids Research 44, W90–W97 (2016).

98. Szklarczyk, D. et al. STRING v11: Protein-protein association networks with increased coverage, supporting functional discovery in genome-wide experimental datasets. Nucleic Acids Res. 47, D607–D613 (2019).

99. Chin, C.-H. et al. cytoHubba: Identifying hub objects and sub-networks from complex interactome. BMC Systems Biology 8, S11 (2014).

100. Shannon, P. et al. Cytoscape: A software environment for integrated models of biomolecular interaction networks. Genome Research 13, 2498–2504 (2003).

101. Freshour, S. L. et al. Integration of the Drug–Gene Interaction Database (DGIdb 4.0) with open crowdsource efforts. Nucleic Acids Research 49, D1144–D1151 (2020).

102. Freshour, S. L. et al. Integration of the Drug-Gene Interaction Database (DGIdb 4.0) with open crowdsource efforts. Nucleic Acids Res 49, D1144–D1151 (2021).

## References

1. Sarma, A., et al. Hyperinflammatory ARDS is characterized by interferon-stimulated gene expression, t-cell activation, and an altered metatranscriptome in tracheal aspirates. bioRxiv (2022).

2. Juss, J. K. et al. Acute respiratory distress syndrome neutrophils have a distinct phenotype and are resistant to phosphoinositide 3-kinase inhibition. Am. J. Respir. Crit. Care Med. 194, 961–973 (2016).

3. Nguyen, E. V. et al. Proteomic profiling of bronchoalveolar lavage fluid in critically ill patients with ventilator-associated pneumonia. PLoS One 8, e58782 (2013).

4. Wang, Z., Beach, D., Su, L., Zhai, R. & Christiani, D. C. A genome-wide expression analysis in blood identifies pre-elafin as a biomarker in ARDS. Am. J. Respir. Cell Mol. Biol. 38, 724–732 (2008).

5. Bhargava, M. et al. Proteomic profiles in acute respiratory distress syndrome differentiates survivors from non-survivors. PLoS One 9, e109713 (2014).

6. Kovach, M. A. et al. Microarray analysis identifies IL-1 receptor type 2 as a novel candidate biomarker in patients with acute respiratory distress syndrome. Respir. Res. 16, 29 (2015).

7. Bhargava, M. et al. Bronchoalveolar lavage fluid protein expression in acute respiratory distress syndrome provides insights into pathways activated in subjects with different outcomes. Sci. Rep. 7, 7464 (2017).

8. Morrell, E. D. et al. Alveolar macrophage transcriptional programs are associated with outcomes in acute respiratory distress syndrome. Am. J. Respir. Crit. Care Med. 200, 732–741 (2019).

9. Christie, J. D. et al. Genome wide association identifies PPFIA1 as a candidate gene for acute lung injury risk following major trauma. PLoS One 7, e28268 (2012).

10. Liao, S. Y. et al. Identification of early and intermediate biomarkers for ARDS mortality by multi-omic approaches. Sci. Rep. 11, 18874 (2021).

11. Jiang, Y., et al. Single cell RNA sequencing identifies an early monocyte gene signature in acute respiratory distress syndrome. JCI Insight 5, (2020).

12. Batra, R. et al. Multi-omic comparative analysis of COVID-19 and bacterial sepsis-induced ARDS. PLoS Pathog. 18, e1010819 (2022).

13. Bime, C. et al. Genome-wide association study in african americans with acute respiratory distress syndrome identifies the selectin P ligand gene as a risk factor. Am. J. Respir. Crit. Care Med. 197, 1421–1432 (2018).

14. Bos, L. D. J. et al. Understanding heterogeneity in biologic phenotypes of acute respiratory distress syndrome by leukocyte expression profiles. Am. J. Respir. Crit. Care Med. 200, 42–50 (2019).

15. Chang, D. W. et al. Proteomic and computational analysis of bronchoalveolar proteins during the course of the acute respiratory distress syndrome. Am. J. Respir. Crit. Care Med. 178, 701–709 (2008).

16. Mirchandani, A. S. et al. Hypoxia shapes the immune landscape in lung injury and promotes the persistence of inflammation. Nat. Immunol. 23, 927–939 (2022).

17. Dong, H. et al. Comparative analysis of the alveolar macrophage proteome in ALI/ARDS patients between the exudative phase and recovery phase. BMC Immunol. 14, 25 (2013).

18. Ren, S. et al. Deleted in malignant brain tumors 1 protein is a potential biomarker of acute respiratory distress syndrome induced by pneumonia. Biochem. Biophys. Res. Commun. 478, 1344–1349 (2016).

19. Tejera, P. et al. Distinct and replicable genetic risk factors for acute respiratory distress syndrome of pulmonary or extrapulmonary origin. J. Med. Genet. 49, 671–680 (2012).

20. Howrylak, J. A. et al. Discovery of the gene signature for acute lung injury in patients with sepsis. Physiol. Genomics 37, 133–139 (2009).

21. Xu, J.-Y. et al. Nucleotide polymorphism in ARDS outcome: A whole exome sequencing association study. Ann. Transl. Med. 9, 780 (2021).

22. Chen, C., Shi, L., Li, Y., Wang, X. & Yang, S. Disease-specific dynamic biomarkers selected by integrating inflammatory mediators with clinical informatics in ARDS patients with severe pneumonia. Cell Biol. Toxicol. 32, 169–184 (2016).

23. Zhang, C. et al. Differential expression profile of plasma exosomal microRNAs in acute type a aortic dissection with acute lung injury. Sci. Rep. 12, 11667 (2022).

24. Kangelaris, K. N. et al. Increased expression of neutrophil-related genes in patients with early sepsis-induced ARDS. Am. J. Physiol. Lung Cell. Mol. Physiol. 308, L1102–13 (2015).

25. Meyer, N. J. et al. IL1RN coding variant is associated with lower risk of acute respiratory distress syndrome and increased plasma IL-1 receptor antagonist. Am. J. Respir. Crit. Care Med. 187, 950–959 (2013).

26. Martucci, G. et al. Identification of a circulating miRNA signature to stratify acute respiratory distress syndrome patients. J. Pers. Med. 11, 15 (2020).

27. Zhu, Z. et al. Whole blood microRNA markers are associated with acute respiratory distress syndrome. Intensive Care Med. Exp. 5, 38 (2017).

28. Englert, J. A. et al. Whole blood RNA sequencing reveals a unique transcriptomic profile in patients with ARDS following hematopoietic stem cell transplantation. Respir. Res. 20, 15 (2019).

29. Lu, X.-G. et al. Circulating miRNAs as biomarkers for severe acute pancreatitis associated with acute lung injury. World J. Gastroenterol. 23, 7440–7449 (2017).

30. Scheller, N. et al. Proviral MicroRNAs detected in extracellular vesicles from bronchoalveolar lavage fluid of patients with influenza virus-induced acute respiratory distress syndrome. J. Infect. Dis. 219, 540–543 (2019).

31. Nick, J. A. et al. Extremes of interferon-stimulated gene expression associate with worse outcomes in the acute respiratory distress syndrome. PLoS One 11, e0162490 (2016).

32. Guillen-Guio, B. et al. Sepsis-associated acute respiratory distress syndrome in individuals of european ancestry: A genome-wide association study. Lancet Respir. Med. 8, 258–266 (2020).

33. Meyer, N. J. et al. ANGPT2 genetic variant is associated with trauma-associated acute lung injury and altered plasma angiopoietin-2 isoform ratio. Am. J. Respir. Crit. Care Med. 183, 1344–1353 (2011).

34. Dolinay, T. et al. Inflammasome-regulated cytokines are critical mediators of acute lung injury. Am. J. Respir. Crit. Care Med. 185, 1225–1234 (2012).

35. Chen, X., Shan, Q., Jiang, L., Zhu, B. & Xi, X. Quantitative proteomic analysis by iTRAQ for identification of candidate biomarkers in plasma from acute respiratory distress syndrome patients. Biochem. Biophys. Res. Commun. 441, 1–6 (2013).

36. Zhang, S. et al. miR-584 and miR-146 are candidate biomarkers for acute respiratory distress syndrome. Exp. Ther. Med. 21, 445 (2021).

37. Shortt, K. et al. Identification of novel single nucleotide polymorphisms associated with acute respiratory distress syndrome by exome-seq. PLoS One 9, e111953 (2014).

38. Bowler, R. P. et al. Proteomic analysis of pulmonary edema fluid and plasma in patients with acute lung injury. Am. J. Physiol. Lung Cell. Mol. Physiol. 286, L1095–104 (2004).

